# Genomic Landscape and Molecular Subtypes of Primary Central Nervous System Lymphoma

**DOI:** 10.1101/2024.10.22.24315961

**Authors:** Shengjie Li, Danhui Li, Zuguang Xia, Jianing Wu, Jun Ren, Yingzhu Li, Jiazhen Cao, Ying Sun, Liyang Zhang, Hongwei Ye, Chengxun Li, Wenjun Cao, Xingtao Zhou, Ying Mao

## Abstract

Primary central nervous system lymphoma (PCNSL) is a rare and aggressive brain tumor with a poor prognosis and is almost exclusively diffuse large B-cell lymphoma (DLBCL). Its genetic characteristics and molecular subtypes in Chinese patients remain poorly understood, which in turn makes developing effective new therapies challenging. By enrolling 176 newly diagnosed Chinese PCNSLs at five tertiary care centers with extensive follow-up, we performed a genomic study aimed at expanding the genomic landscape and developing new molecular subtypes. We first confirmed that the molecular subtyping of DLBCL, as previously published, is not applicable to Chinese PCNSLs. We then identified (n = 58) and validated (n = 82) three prominent genetic subtypes related to different clinical and molecular features of PCNSL, and further confirmed by an independent external Chinese PCNSL cohort (n = 36). We called these BMIs (from the co-occurrence of mutations in two genes among BTG1, MYD88, and IRF4), which are associated with favorable outcomes; E3s (so-called EP300 mutations), which are associated with unfavorable outcomes; and UCs (unclassified, without characteristic mutations). Importantly, EP300 was mutated in more Asians (16.98%) than in Western PCNSLs (< 4.53%), resulting in unfavorable outcomes independent of the specific mutation site. Our analysis comprehensively reveals the genomic landscape of Chinese PCNSL and emphasizes the clinical value of molecular classification for improving precision medicine strategies.

## Introduction

Primary central nervous system lymphoma (PCNSL), a rare subtype of extranodal non-Hodgkin lymphoma, accounts for >60% of PCNSL cases and has an unfavorable prognosis even when the patient receives standard treatment, which is based on a high-dose methotrexate (HD-MTX) regimen^1,2^. PCNSL is a unique lymphoma that differs from other tumors and is characterized as a genetically heterogeneous disorder marked by a variety of low-frequency mutations, somatic copy number alterations, and structural variants^3–6^. Thus, combining standard immunochemotherapy with promising novel agents that target specific pathways via different molecular clusters may improve patient prognosis^7,8^.

Pathologically, nearly 95% of PCNSLs are diffuse large B-cell lymphomas (DLBCLs). Recently, DLBCL has been categorized into various molecular clusters on the basis of genomic sequencing and RNA sequencing, including quartet categorization by *L.M. Staudt*’s team^9^, quintet categorization by *M.A. Shipp*’s team^10^, *L.M. Staudt*’s team’s refined septet method^11^, and the LymphPlex classification proposed by *W.L. Zhao*’s team^12^. However, PCNSL has been proven to be a biological entity that is molecularly distinct from DLBCL^13^, so these molecular clusters of DLBCL may not be applicable to PCNSL. Notably, the integration of genome-wide data from multiomic studies by *A. Alentorn* et al.^14^ showed four molecular patterns of PCNSL mainly use gene expression data. These patterns have a distinctive prognostic impact, providing a basis for future clinical stratification and subtype-based targeted interventions. Owing to the molecular heterogeneity between various ethnic groups of PCNSL patients^3,15^, the applicability of the proposed classifications for PCNSL to other populations requires further confirmation. No effective biomarker or molecular classification scheme exists to tailor therapies for individual PCNSL patients. Furthermore, the issue of PCNSL molecular heterogeneity in Chinese patients has not been adequately addressed, primarily because the data have come from small, single-center studies of Chinese PCNSL patients^16,17^.

To address these issues, we conducted whole-exome sequencing (WES) on specimens obtained from 140 patients with newly diagnosed PCNSL who were seen at one of three tertiary care cancer centers. These patients were divided into a discovery cohort and a validation cohort. Among the 140 patients, 94.29% received treatment with an HD-MTX-based chemotherapy regimen followed by consolidation therapy (detailed in Table S1), and all 140 patients underwent long-term follow-up. Additionally, WES/Whole Genome Sequencing (WGS) data were obtained from 36 Chinese PCNSL patients at two other tertiary care cancer centers^17^, serving as an independent external cohort. This representative and clinically annotated PCNSL cohort was utilized to comprehensively detect mutations. Importantly, to facilitate routine clinical implementation, we identified three molecular subtypes composed only of single-nucleotide variations (SNVs); these SNVs were associated with different outcomes, and this was confirmed in the validation cohort and in an independent external Chinese PCNSL cohort (n=36)^17^. Our results comprehensively reveal the genomic landscape of PCNSLs in Chinese patients and provide a clinically actionable PCNSL classification system, thereby improving precision medicine strategies.

## METHODS

### Sample collection and clinicopathological features of PCNSL patients

In this study, a total of 140 newly diagnosed PCNSL patients were retrospectively enrolled from January 2010 to December 2020 from three independent medical centers, namely, Huashan Hospital of Fudan University, Renji Hospital of Shanghai Jiao Tong University, and Shanghai Cancer Center of Fudan University. The study comprised two cohorts: the discovery cohort, which included 58 patients from Huashan Hospital of Fudan University, Shanghai; and the validation cohort (n = 82), which consisted of 25 patients from Huashan Hospital of Fudan University, 51 patients from Renji Hospital of Shanghai Jiao Tong University, and 6 patients from Shanghai Cancer Center of Fudan University. In the discovery cohort, paired blood samples were also obtained for analysis in conjunction with each tumor sample. In the validation cohort, only unpaired FFPE tumor tissue samples were obtained for analysis. Additionally, WES/WGS data of 36 patients newly diagnosed with PCNSL from Fujian Cancer Hospital and The First Affiliated Hospital of Fujian Medical University between February 2012 and October 2020 were included in this study, as an independent external Chinese PCNSL cohort. The general clinical characteristics of these 36 PCNSL patients and the sequencing protocol have been previously described^17^.

None of the patients had received steroid treatment for PCNSL, and PCNSL tumors were diagnosed on the basis of the World Health Organization criteria^13^. All patients underwent a 2-deoxy-2[F-18] fluoro-D-glucose positron emission tomography/computed tomography (FDG PET/CT) scan and bone marrow aspiration to exclude systemic tumor manifestation on the basis of the guidelines of the International PCNSL Collaborative Group^18^ and the European Association of Neuro-Oncology^19^. PCNSL patients for whom ocular involvement was present were excluded. Each tumor tissue sample was independently reviewed by at least 2 pathologists to confirm that the tumor sample was histologically consistent with the PCNSL. All patients underwent long-term follow-up, which ran to December 30, 2022.

### Ethics statement

This study was conducted with the approval of the Institutional Review Boards at Huashan Hospital of Fudan University (Approval No. 2022-529), Shanghai Cancer Center of Fudan University (Approval No. 1612167-18), Renji Hospital of Shanghai Jiao Tong University (Approval No. LY2024-112-C), and Fujian Cancer Hospital (Approval no. SQ 2020-106-01). All participants provided written informed consent before participating in the clinical study and before the collection of tumor tissues. The research was carried out in strict adherence to the ethical principles of the Declaration of Helsinki and conformed to international norms of good clinical practice.

### Treatment Regimens

All PCNSL patients were treated with HD-MTX-based combination immunochemotherapy, followed by consolidation therapy (detailed in Table S1). The following chemotherapy protocols were used for induction: HD-MTX combined with IDA, HD-MTX combined with R, HD-MTX combined with IDA and R, or a combination of BTK inhibitors. Whole-brain radiation therapy or stem cell transplantation is a form of consolidated therapy. The detailed therapeutic schedule was the same as that previously described^20^.

### Sample processing

The collection and processing of samples for this study were carried out by the Department of Pathology or Department of Neurosurgery at Huashan Hospital of Fudan University, Renji Hospital of Shanghai Jiao Tong University, and Shanghai Cancer Center of Fudan University. In the discovery cohort, surplus tumor tissues and paired blood samples obtained during surgical procedures were collected. To guarantee the quality of these samples, each specimen was promptly and accurately labeled with relevant patient information within 30 minutes of collection and then immediately snap-frozen in liquid nitrogen. These samples were stored at -80°C until being shipped to GenomiCare Biotechnology (Shanghai, China). In the validation cohort, the residual tumor tissue specimens were subjected to FFPE. These FFPE samples were transported in a dry ice container to Sinotech Genomics Technologies in Shanghai, China, accompanied by a time and temperature tracker to ensure proper monitoring during shipment. Upon arrival, the samples were promptly stored in liquid nitrogen to maintain their integrity until further processing.

### DNA extraction

For frozen fresh tissue and matched peripheral blood samples, total genomic DNA (gDNA) was extracted via the Maxwell RSC Blood DNA Kit (AS1400, Promega) on a Maxwell RSC system (AS4500, Promega) following the manufacturer’s instructions. For FFPE tissue, total gDNA was extracted via the QIAamp DNA FFPE Tissue Kit (56404, Qiagen) following the manufacturer’s instructions. The integrity and concentration of the gDNA were determined via the Qsep100 System (BIOptic, China) and a Qubit 3.0 fluorometer (Thermo Fisher Scientific). The OD_260_ was measured with a NanoDrop One (Thermo Fisher Scientific).

### WES library preparation and sequencing

For frozen tissue and matched peripheral blood samples, exome DNA was extracted via the SureSelect Human All Exon V7 Kit (5991-9039EN, Agilent). The library was subsequently prepared via the SureSelectXT Low Input Target Enrichment and Library Preparation system (G9703-90000, Agilent). For the FFPE sample, the exome DNA was captured via the SureSelect Human All Exon V8 Kit (5191-6874, Agilent) and prepared into a library via the SureSelect XT HS2 DNA Reagent Kit (G9983A, Agilent). The library was validated via the use of an Agilent 2100 Bioanalyzer and a Qubit 3.0 fluorometer. Paired-end 150 bp read sequencing was performed on an Illumina NovaSeq 6000. Image analysis and base calling were performed via onboard RTA3 software (Illumina).

### Somatic mutation calling

Quality control was conducted via FastQC software. The cleaned and trimmed FASTQ files were aligned to the UCSC human reference genome (hg19) via the Burrows–Wheeler Aligner (BWA) with default parameters. For each paired blood sample, SNVs and short insertions/deletions (INDELs) were identified via Sentieon TNseq^21^ with default parameters. The identified somatic mutations were removed if they did not satisfy any one of the following criteria: a variant allele frequency (VAF) of at least 0.05, support from a minimum of three reads, annotation by the Variant Effect Predictor (VEP) package^22^, and transformation into a mutation annotation format (MAF) file for further analysis with maftools^23^.

When a paired normal sample (FFPE sample) was not available, a panel of normal (PON) files was used as a paired control to call the SNVs and InDels. A panel of normal files was created using the reads from clinical blood samples that showed no evidence of tumor contamination from 100 different individuals collected by Genomicare Biotechnology (Shanghai). The reads were retained when they passed the standard WES QC, as described in the WES subsection. The reads for reference alleles and alternative alleles were gathered for each candidate site in all 100 samples, except for false positives, which were those exhibiting a candidate allele frequency exceeding 0.05 in more than 5 samples.

We applied an additional PON mask for all the candidate somatic SNV and INDEL sites to exclude germline mutations, as previously reported^10,24^. In general, in addition to the standard retention rules, mutations were retained if they were known driver alterations highlighted as being biologically significant in the COSMIC database. Mutations with a tumor mutation frequency ≥20% were also retained. Other germline mutations were retained if their existence ratio in the Exome Aggregation Consortium (ExAC) exac_all was < 0.00005, their existence ratio in the Asian population exac_eas was < 0.0003, and their existence ratio in previous GenomiCare samples germline_gc was < 0.0006. Retained mutations were considered somatic mutations.

Thus, the PON file and PON mask limit possible recurrent artifacts of sequencing and the presence of germline mutations in somatic mutations that are falsely selected owing to a lack of paired blood data.

### Somatic copy number alteration calling and profiling

Using the method described in the ExomeCNV package^25^, we implemented a normalized depth‒coverage ratio method to identify CNVs in paired samples. To correct for five potential biases (the size of exonic regions, batch effects, both the quantity and quality of sequencing data, local GC content, and genomic mappability) that affect the raw read counts, we utilized a standard normal distribution model. Genes with a haploid copy number ≥3 or ≤1.2 were defined as amplified or deleted, respectively, and a minimum tumor content (purity) of 20% was needed.

### Significance analysis of recurrent somatic copy number alterations

Sequenza^26^ was used to generate the segment files. The segment files were used as the input for GISTIC2.0^27^ to detect recurrent arm-level and focal peaks in copy number alterations via the parameters -js 40 -conf 0.99 -ta 0.2 -td 0.2 -brlen 0.8, with other parameters set to the defaults.

### Cancer-driver gene mutations filtering

After the aforementioned steps of calling SNVs and CNVs, we further filtered the resulting mutated genes by intersecting them with a designated group of genes recognized as cancer-related genes. This group of genes was sourced from two prominent public cancer gene databases. Part 1 of the group was obtained from the OncoKB curated cancer gene list (https://www.oncokb.org/cancerGenes)^28^. These genes are classified as cancer genes by OncoKB on the basis of their inclusion in various sequencing panels, the Sanger Cancer Gene Census, or the criteria established by *Vogelstein* et al.^29^. Part 2 was derived from the ranked CIViC gene candidates table (https://github.com/griffithlab/civic-server/blob/master/public/downloads/RankedCivicGeneCandidates.tsv)^30^. The final list of cancer-related genes consisted of all genes in Part 1 and genes in Part 2 only when its ‘panel_count’ value (in the CIViC tsv file listed above) was ≥ 2.

### Bioinformatic analysis

The mutational signature classification was based on all single-base substitution (SBS) signatures of the COSMIC mutational signatures^31^. TMB was defined as the aggregate count of somatic nonsynonymous mutations, including SNVs or INDELs, within the tumor exome for each patient^32^. This count was then divided by the overall size of the targeted regions (35 for WES), resulting in the TMB, expressed in counts per megabase (counts/Mb). The MATH score^33^ was calculated on the basis of the width of the variant allele frequency (VAF) distribution, utilizing maftools for the analysis^23^. The pathway map was generated via maftools as previously described^34^.

### Classification model for identifying molecular types

CoxNet survival analyses and least absolute shrinkage and selection operator (LASSO) regressions were performed via the scikit-learn package in Python (version 3.9.7). K‒M survival curves (log-rank test) were used for OS analysis via the survival and survminer packages in R (version 4.1.2).

#### (1) Selection of 24 initial features in the discovery cohort

Molecular features derived from SNVs were selected to develop subsequent classification models.

First, to identify genes with high variability for use in clustering, those with low mutation frequencies were filtered out. This process led to the selection of genes with a mutation frequency greater than 5% for clustering purposes. A total of 150 genes were screened for further analysis.

In the second phase, OS was analyzed as the dependent variable, with the aforementioned 150 genes serving as independent variables. The feature selection steps were as follows: (1) a comprehensive approach involving CoxNet survival analyses and LASSO regressions was employed to identify characteristic genes; (2) the importance score of each feature in the model was calculated and ranked in descending order; (3) features that ranked in the top 50% were recorded as potential features once; (4) processes 1-3 were repeated 500 times, and all potential features were recorded; and (5) among the feature sets obtained in the above process, those recorded more than 250 times were selected.

Ultimately, 24 feature genes, namely, EP300, KMT2D, MPEG1, TUSC3, MYD88, PTCH1, DST, ZNF521, BTG1, NOTCH2, PRDM1, ADGRL3, CDH11, CREBBP, ATRX, TCL1A, GNA13, ETV6, IRF4, HIST2H3D, CD79B, GRM8, MYH11, and SPTA1, were selected.

#### (2) Differential feature selection

To determine the most suitable features among the initial 24 for the classification model, Kaplan‒Meier analyses were conducted. Features qualifying for further analysis met the following criteria: a log-rank test P value of less than 0.2 and any group with more than 5 patients. Ultimately, 6 DEGs, namely, EP300, CREBBP, DST, MYD88, BTG1, and IRF4, were identified.

#### (3) Model training utilizing random combinations of 6 differential features

To adhere to clinical practice guidelines for predicting patient outcomes and implementing individualized management strategies for PCNSL patients, patients with PCNSL were categorized into three subgroups (those with good, poor, and intermediate prognoses) in the present study. Six differential features were ultimately selected for building a classification model to classify patients into the three subtypes. These features were randomly combined to form three types, yielding a total of 288 combinations. To identify the most appropriate classification model among these combinations, Kaplan‒Meier analyses were performed. Models were chosen on the basis of a log-rank test P value of less than 0.05 and subgroup sample sizes greater than 8. Ultimately, seven candidate molecular subtype combinations met these criteria: (molecular markers shown in the format of subtype 1 vs. subtype 2 vs. subtype 3): ① EP300 (predominant) vs. MYD88/BTG1/IRF4≥2 vs. others; ② EP300 vs. MYD88/BTG1/IRF4≥2 (predominant) vs. others; ③ BTG1 (predominant) vs. CREBP/DST≥1 vs. others; ④ BTG1 vs. CREBP/DST≥1 (predominant) vs. others; ⑤ BTG1/IRF4≥1 vs. EP300/CREBP/DST≥1 vs. others; ⑥ BTG1/IRF4≥1 vs. EP300/DST≥1 vs. others; and ⑦ CREBP/DST≥1 vs. MYD88/BTG1/IRF4≥2 vs. others.

#### (4) Validation of candidate molecular subtypes

The WES data and OS data of 82 patients in the validation cohort were included to validate the candidate molecular subtypes. The mutation frequencies of EP300, CREBBP, DST, MYD88, BTG1, and IRF4 in the validation cohort were greater than 5%. Kaplan–Meier survival analysis was used to validate the seven candidate molecular subtype combinations. The performance of [EP300 (predominant) vs. MYD88/BTG1/IRF4≥2 vs. others] (3-way P = 0. 0202) and [EP300 vs. MYD88/BTG1/IRF4≥2 vs. others (predominant)] (3-way P = 0. 0346) was better than those of the other five molecular subtypes (3-way P > 0.05 for both). Additionally, compared with the [EP300 vs. MYD88/BTG1/IRF4≥2 vs. others (predominant)] molecular subtype, the Kaplan–Meier survival curve of [EP300 (predominant) vs. MYD88/BTG1/IRF4≥2 vs. others] showed no crossover. Finally, the performance of [EP300 (predominant) vs. MYD88/BTG1/IRF4≥2 vs. others] was the best, so we selected it as the molecular subtype of Chinese PCNSL patients. In other words, PCNSL patients with both EP300 and BMI (BTG1, IRF4, and MYD88) variants were categorized into the E3 subtype.

### Hematoxylin & eosin and immunohistochemical staining

PCNSL tumor tissue was fixed with 10% paraformaldehyde and embedded in paraffin. The paraffin-embedded tissues were cut into 4-µm-thick sections. These sections were then subjected to staining with hematoxylin and eosin (H&E) or for immunohistochemistry. For H&E staining, the sections were stained with hematoxylin and eosin following standard protocols (ab245880, Abcam). Immunohistochemistry was performed for ki-67 (ab15580, Abcam), CD2 (ab314761, Abcam), CD5 (ab75877, Abcam), CD10 (ab256494, Abcam), CD19 (ab134114, Abcam), CD20 (ab78237, Abcam), CD79a (ab79414, Abcam), BCL2 (MA5-11757, Thermo Scientific), BCL6 (PA5-14259, Thermo Scientific), MUM1 (ab247079, Abcam), c-Myc (MA1-980, Thermo Scientific), and p53 (MA5-14516, Thermo Scientific) via an automated Leica BOND-III staining system. H&E and immunohistochemistry images were obtained with a KFBIO scanner (KF-PRO-005-EX, Zhejiang, China) and visualized via KFBIO SlideViewer software. Each staining was independently reviewed by at least 2 pathologists.

### In situ hybridization of fluorescence

PCNSL tumor tissue was fixed with 10% paraformaldehyde and embedded in paraffin. The paraffin-embedded tissues were cut into 4-µm-thick sections. Fluorescence in situ hybridization was performed in accordance with standard protocols via the following commercial probes: MYC break-apart, BCL-2 break-apart, and BCL-6 break-apart probes (LBP Medicine Science & Technology Co., Ltd., Guangzhou, China). Staining was independently evaluated by two hematopathologists, and discrepancies were resolved by another hematopathologist.

### HIV, HBV, and EBV detection

Antibody and antigen levels of HIV were measured via an Elecscy HIV combi PT assay kit

(Roche, Germany) with a Cobas e 601 analyzer (Roche). Hepatitis B virus (HBV)-specific PCR was performed via an HBV virus nucleic acid test kit (Sansure Biotech, China) with an Applied Biosystems 7500 Real-Time PCR System (Thermo Scientific). EBV-encoded RNA in situ hybridization (EBER-ISH) was performed to assess the presence of Epstein-Barr virus (EBV) in tumor tissue. The testing utilized a fluorescein isothiocyanate -coupled specific peptide nucleic acid probe (Roche Diagnostics, Mannheim, Germany).

### Statistical analysis

All the statistical analyses were performed via R version 4.1.2 and GraphPad Prism version 9. The specific statistical analyses employed for each figure are detailed in the respective legends. We used PASS software to calculate the sample size for our study, with a significance level of 0.05 and a power of 0.80. The observed group proportions were 0.6, 0.2, and 0.2, whereas the expected proportions were 0.33 for each group. The sample size required to meet these criteria was 25 participants. Consequently, the sample size utilized in this study is deemed adequate. Whole-exome sequencing data were processed via nf-core/sarek v3.4.2^35^ of the nf-core collection of workflows^36^, which uses reproducible software environments from the Bioconda^37^ and Biocontainers^38^ projects. The pipeline was executed with Nextflow v24.04.2^39^.

## Results

### PCNSL genomic landscape

Using WES, we detected mutations in 140 patients who were newly diagnosed with PCNSL at one of three cancer centers; in 59% of cases, blood samples were lacking. Because EBV-positive PCNSL is recognized as an immunodeficiency-associated PCNSL in the current WHO CNS5 classification^40,41^, all participants in the study who tested negative for EBV were included. Additionally, DLBCL patients with HBV and HIV infection may have different mutational profiles^42,43^; thus, all participants in the study who tested negative for HIV and HBV were included. The key demographic and clinical characteristics of the patients are summarized in **Figure S1** and **Table S1**. The study design and flow chart are displayed in **Figure 1A**. After filtering, we identified 115,938 genetic variation events in the 140 PCNSL samples analyzed (median = 19.06 mutations/Mb, range = 0.24–91.2 mutations/Mb; median number of variants = 949.5/sample) (**Figure S2A, Figure S3A**).

**Figure 1.**
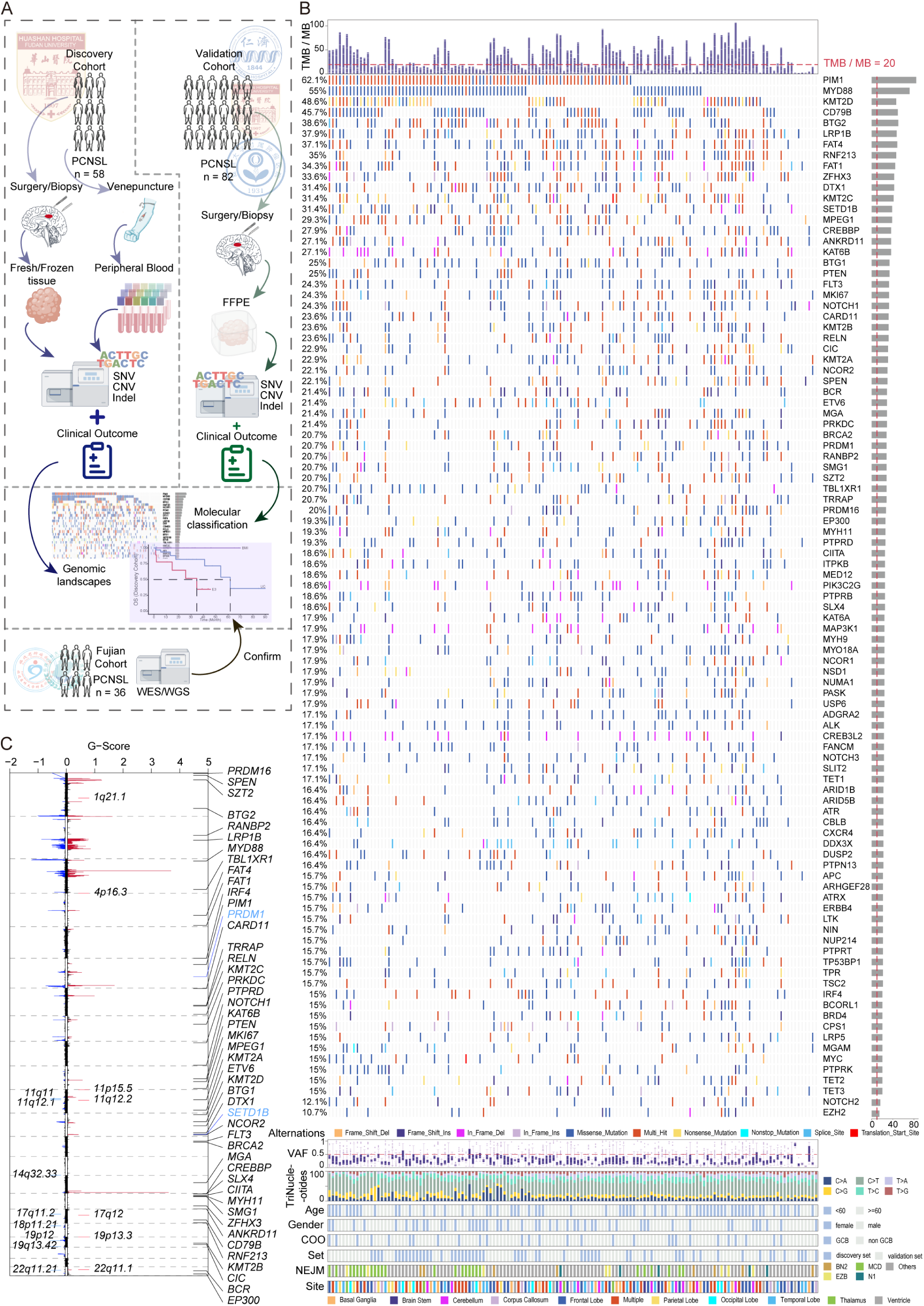
Study design and the PCNSL mutational landscape in Chinese patients. A: Study design, including workflow and data composition for the discovery cohort and validation cohort. B: Number and frequency of recurrent mutations. Gene‒sample matrix of recurrently mutated genes ranked by mutation frequency (n = 140). The total mutation density across the cohort is displayed at the top, with the variant allele fraction and trinucleotides at the bottom. C: GISTIC2.0 results of significant recurrent focal amplifications (red) and deletions (blue). Genes affected by each focal event are annotated (n = 140). X-axis: plot of chromosomes; Y-axis: G score.

We applied a filter to the 140 PCNSLs on the basis of the genes curated in the CIViC and OncoKB databases to identify candidate cancer genes with hallmark mutations in PCNSL. Some of the mutated genes were PIM1 (in 62.1% of PCNSLs), MYD88 (55.0%), KMT2D (48.6%), CD79B (45.7%), BTG2 (38.6%), LRP1B (37.9%), FAT4 (37.1%), RNF213 (35%), FAT1 (34.3%), ZFHX3 (33.6%), DTX1 (31.4%), KMT2C (31.4%), SETD1B (31.4%), MPEG1 (29.3%), CREBBP (27.9%), ANKRD11 (27.1%), KAT6B (27.1%), BTG1 (25%), and PTEN (25%) (**Figure 1B**), which are involved in chromatin histone modification, BCR-TLR-mediated NF-κB signaling, immune signaling, the cell cycle, PI3 kinase signaling, MAPK signaling, and Wnt/β-catenin signaling (**Figure S2B**)^3,5,9–11,44–51^. In this study, we also identified several other pathways in PCNSL, such as pathways related to genome integrity, RTK signaling, RNA abundance, TGFB signaling, TOR signaling, and apoptosis (**Figure S2B**), many of which have defined roles in other cancers^34^. The mutually exclusive or cooccurring relationships of these genes are shown in **Figure S3D**. Survival associated with candidate cancer genes with mutation frequencies greater than 15% plus the NOTCH2 and EZH2 genes^9^ is shown in **Figure S4**.

In the search for focal copy number alterations (CNAs) in the 140 PCNSLs, we detected significant recurrent amplifications at chromosomal locations 1q21.1, 4q16.3, 11p15.5, 11q12.2, 17q12, 19p13.3, 22q11.1, Yq11.221, and Yq12. We also detected deletions at 11q11, 11q12.1, 14q32.33, 17q11.2, 18p11.21, 19p12, 19q13.42, and 22q11.21 (**Figure 1C**). The arm-level CNAs among all 140 PCNSL samples are displayed in **Figure S2C**. The pattern of somatic mutations caused by the different mutational processes in the genome, termed *mutational signatures*, was calculated and compared to the well-established signatures in COSMIC^52^. The mutational signature contributions are shown in **Figure S2D**. The COSMIC 5, COSMIC 45, and COSMIC 1 signatures contributed the most to the PCNSL genome (**Figure S2E**). These factors are associated with aging, tobacco smoking, NER deficiency, the regulation of oxidative DNA damage repair, and the spontaneous deamination of 5-methylcytosine. Taken together, these results establish a comprehensive genomic landscape of PCNSL in Chinese individuals.

### Comparison of the genomic landscapes of the discovery and validation cohorts

Owing to differences in sample type between the discovery and validation cohorts (fresh tumor [discovery cohort] vs. formalin-fixed paraffin-embedded [FFPE] [validation cohort]) and the presence or absence of matched samples (paired [discovery cohort] vs. tumor-only [validation cohort]), we applied further filtering and recalibration steps to reduce false-positive calls in the validation cohort (for details, see the Methods subsection on somatic mutation calling) and compared the genomic landscapes of the discovery and validation cohorts. The key demographic and clinical characteristics of the patients in the discovery and validation cohorts are summarized in **Table S2**. We identified 12,222 genetic variation events (median = 3.84 mutations/Mb, median variants = 159/sample, **Figure S3B, Figure S5A**) in the analyzed discovery cohort of PCNSL samples and 103,716 genetic variation events (median = 6.85 mutations/Mb, median variants = 1597/sample, **Figure S3C, Figure S5B**) in the analyzed validation cohort of PCNSL samples. The genetic variation profiles of the discovery and validation cohort of PCNSL samples were similar, as revealed by principal component analysis (PCA) (**Figure 2A**), but the mutation rate was higher in the validation cohort of PCNSL samples (**Figure 2B**).

**Figure 2.**
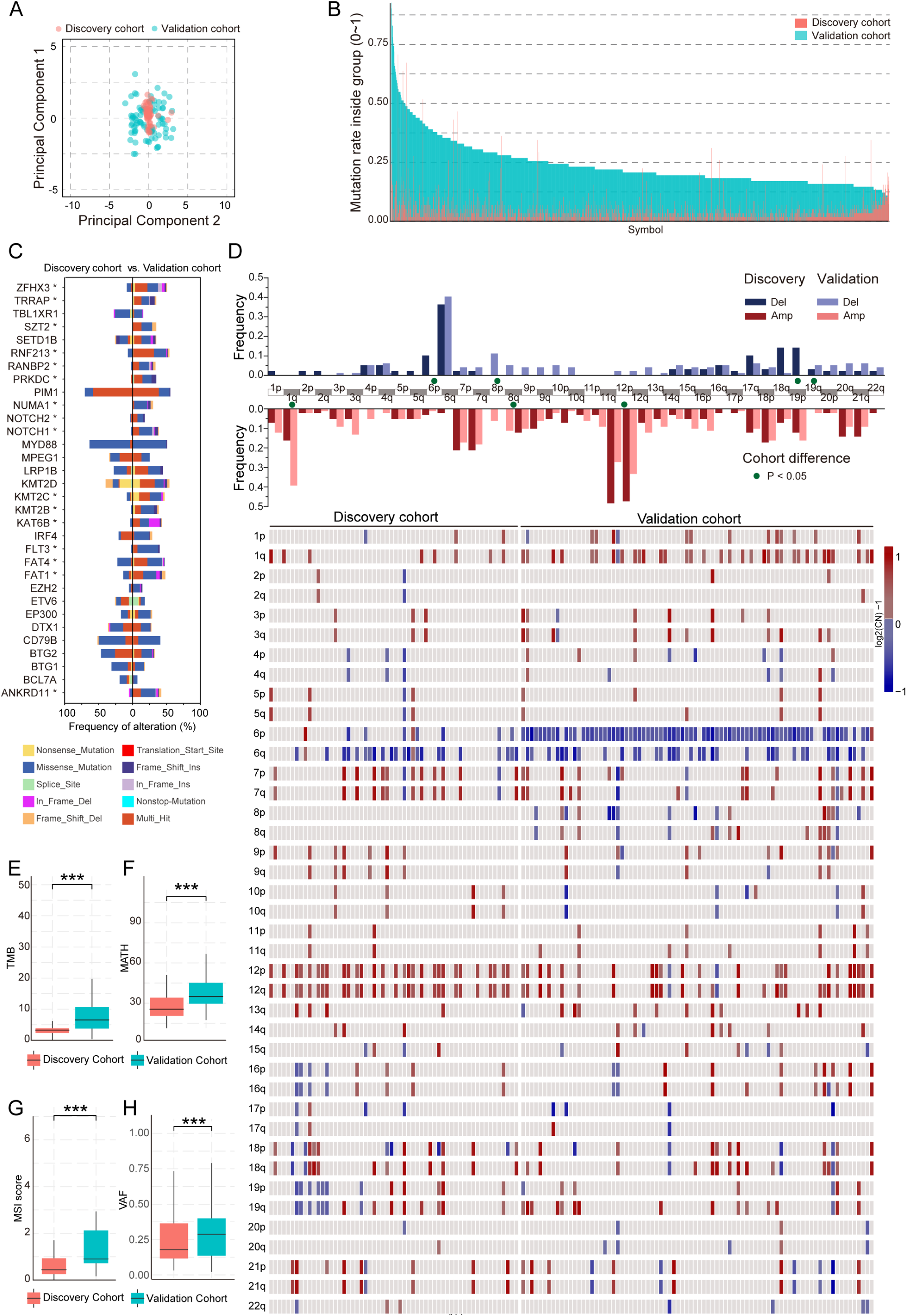
Comparison of mutation profiles between the discovery cohort and validation cohort of PCNSL patients. A: Principal component analysis of whole-exome sequencing (WES) data between the discovery cohort and validation cohort of PCNSL patients B: Mutation rate of WES data between the discovery cohort and validation cohort of PCNSL patients C: A mirror bar plot showing the frequencies of genetic alterations in the discovery cohort of PCNSL patients compared with those in the validation cohort. D: Arm-level copy number alterations of the discovery cohort and validation cohort samples are displayed. The frequencies of amplifications and deletions were compared. E: Comparison of the tumor mutational burden (TMB) between the discovery cohort and the validation cohort of PCNSL patients F: The levels of mutant-allele tumor heterogeneity (MATH) in the discovery cohort and the validation cohort of PCNSL patients were compared. G: The levels of microsatellite instability (MSI) in the discovery cohort and the validation cohort of PCNSL patients were compared. H: The variant allele frequency (VAF) in the discovery cohort and the validation cohort of PCNSL patients were compared. The discovery cohort included blood-paired fresh tumor tissue samples (n = 58), and the validation cohort included unpaired FFPE tumor tissue samples (n = 82). Independent-samples t tests and chi-squared tests were used. \**P* < 0.05; \*\**P* < 0.01; \*\*\**P* < 0.001.

PIM1, MYD88, CD79B, and KMT2D were the most frequently mutated candidate cancer genes of PCNSL. PIM1 (70.7%), MYD88 (63.8%), CD79B (51.7%), and KMT2D (39.7%) (**Figure S5C**) were identified in the discovery cohort, and PIM1 (56.1%), MYD88 (48.8%), CD79B (41.5%), and KMT2D (54.6%) were also observed at similar mutation frequencies in the validation cohort (**Figure S5D**). However, several of the less frequently mutated candidate cancer genes in PCNSL whose mutation frequencies were significantly different when the discovery and validation cohorts were compared included ZFHX3, TRRAP, SZT2, RNF213, RANBP2, PRKDC, NUMA1, NOTCH1, NOTCH2, KMT2C, KMT2B, KAT6B, FLT3, FAT4, FAT1, and ANKRD11 (all *P* < 0.05) (**Figure 2C**). The arm-level CNAs between the discovery and validation cohort of PCNSL samples were mostly similar and are displayed in **Figure 2D**. Significant differences in both recurrent amplifications (1q, 8q, and 12p) and deletions (6p, 8p, 19p, and 19q) were observed. The results of significant recurrent focal amplifications and deletions were visualized via GISTIC 2.0 in the discovery (**Figure S5E**) and validation (**Figure S5F**) cohorts.

The tumor mutational burden (TMB) describes the number of mutations in one tumor sample and is often related to the prognosis of patients receiving clinical treatments^53^. The mutant-allele tumor heterogeneity (MATH) score reflects intratumor heterogeneity^33^. Microsatellite instability (MSI) arises from the impaired function of DNA mismatch repair proteins, and its molecular characteristics are regarded as significant genetic markers^54^. Although the TMB (**Figure 2E**), MATH score (**Figure 2F**), MSI (**Figure 2G**), and the variant allele frequency (VAF) (**Figure 2H**) were greater in the validation cohort than in the discovery cohort (*P* < 0.001), the mean values were relatively close. The 3 signatures extracted from the discovery samples presented cosine similarities of 86.7%, 93.1% and 94.4% to COSMIC signatures 6, 45, and 10a, respectively (**Figure S5G**), whereas those extracted from the validation samples presented similarities of 80.4%, 70.9%, and 77.9% to COSMIC signatures 5, 1, and 23, respectively (**Figure S5H**). Taken together, these results suggest that the genetic variation profiles of the discovery and validation PCNSL samples exhibit certain discrepancies.

### The unique genomic landscape of PCNSLs in Chinese patients

To date, only single-center studies with small sample sizes have reported heterogeneity in the genomic landscape among different populations of Chinese PCNSL patients^15^. Thus, in this large, multicenter study, we further compared the genetic landscape of our cohort with previously published PCNSL/DLBCL data. The frequencies of recurrent genetic alterations in PCNSL patients were compared with those in DLBCL patients from the United States, Finland, Denmark, Israel, the United Kingdom, Hong Kong, and India^49^. Mirror bar plots (**Figure 3A**) revealed that a set of genes, such as PIM1 (62.1% vs. 16.88%), MYD88 (55% vs. 18.01%), IRF4 (15% vs. 3.98%), and EP300 (19.29% vs. 5.97%), had significantly higher mutation rates in Chinese PCNSL patients than in DLBCL patients, suggesting genetic diversity of the cancer genome between diseases. Compared with those in PCNSLs in French patients ^14^ (**Figure 3B**), the mutation frequencies of EP300 (19.29% vs. 3.48%) and KMT2D (48.6% vs. 22.61%) were significantly higher in the PCNSLs of Chinese patients. Compared with those in the PCNSLs^3^ of Japanese patients (**Figure 3C**), the mutation frequencies of MYD88 (55% vs. 85.6%) and BTG2 (38.6% vs. 87.8%) were significantly lower in the PCNSLs of Chinese patients. These data revealed a unique mutational landscape of Chinese PCNSL patients in terms of the frequency of SNVs. Next, the frequencies of recurrent SNAs with the GISTIC2.0 program in Chinese PCNSL patients were compared with those in DLBCL patients. At the focal level of SNAs, Chinese PCNSL samples had significant levels of 11q12.4, 18p11.21, 19q13.42, 22q11.1, and 22q11.21 deletions along with amplifications in arms 1q21.1, 4p16.3, 11q12.2, and 17q12 (**Figure 3D**). Compared with PCNSLs from Japanese patients, those from Chinese patients also had significant levels of deletions in arms 11q12.1, 18p11.21, 19q13.42, 22q11.1, and 22q11.21 along with amplifications in arms 1q21.1, 4p16.3, 11q12.2, and 17q12 (**Figure 3E**). These results suggest genetic diversity in the PCNSL genome between races.

**Figure 3.**
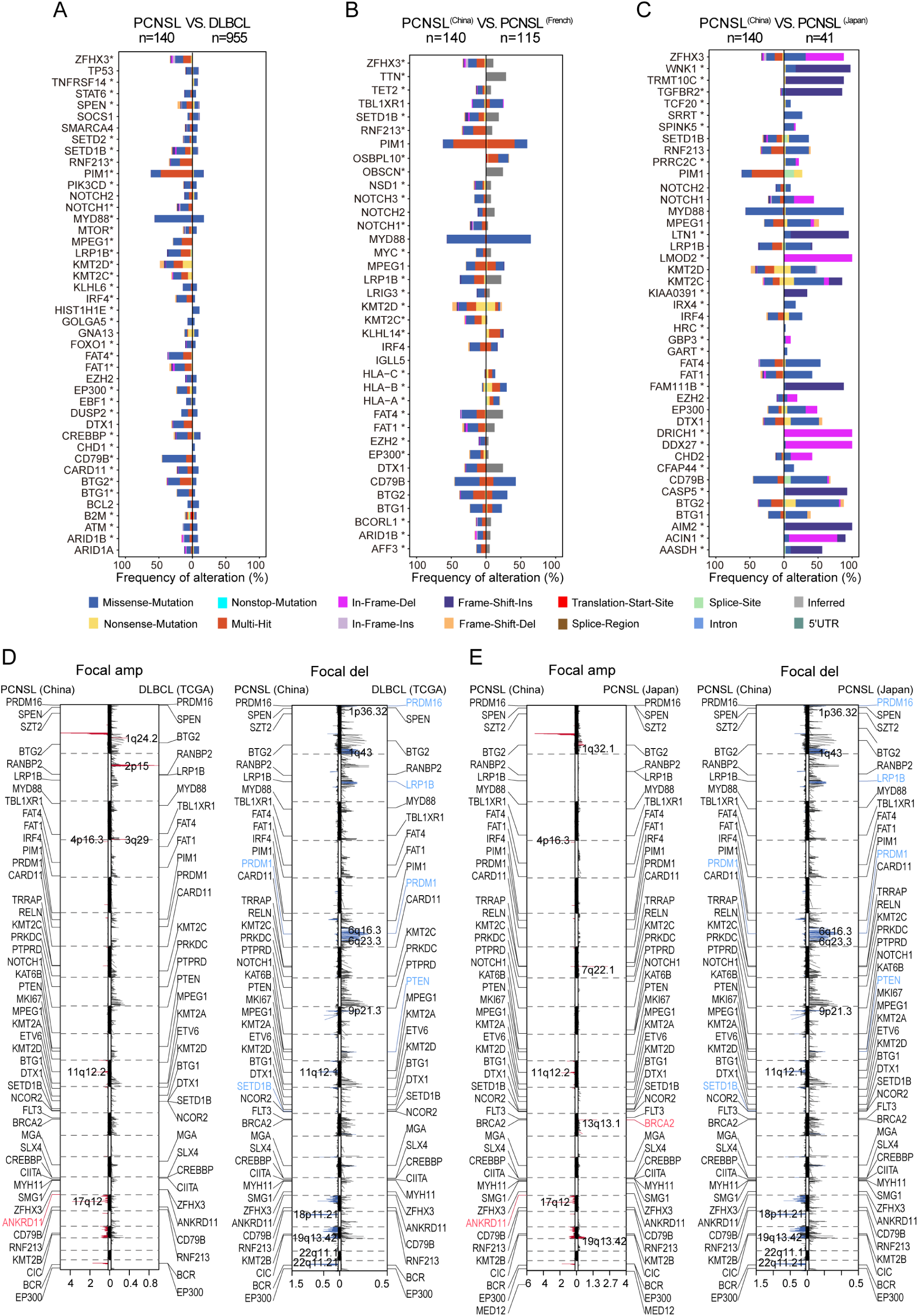
Mutation profiles of Chinese PCNSLs, DLBCLs, and PCNSLs of other races. A: A mirror bar plot showing the frequencies of genetic alterations between Chinese PCNSL patients (n = 140) and DLBCL patients (n = 955). B: A mirror bar plot showing the frequencies of genetic alterations between Chinese PCNSL patients (n = 140) and French PCNSL patients (n = 115). C: A mirror bar plot showing the frequencies of genetic alterations between Chinese PCNSL patients (n = 140) and Japanese PCNSL patients (n = 41). D: GISTIC-defined recurrent copy number focal deletions (blue) and gains (red) as mirror plots in DLBCL (TCGA, n = 46) and Chinese PCNSLs (n = 140) are shown. X-axis: plot of chromosomes; Y-axis: G score. E: The GISTIC2.0-defined recurrent copy number focal deletions (blue) and gains (red) in Japanese PCNSL patients (n = 41) and Chinese PCNSL patients (n = 140) are shown as mirror plots. Y-axis: plot of chromosomes; X-axis: G score. The chi-squared test was used. \**P* < 0.05. In the figure legend of A-C, ‘Inferred’ means that the mutation types were not directly provided but were calculated from the supplemental tables of the corresponding paper; thus, the results are ‘Inferred’, and the *in facto* mutation number will not be less than the inferred value.

Owing to the genetic diversity of the PCNSL genome across races and the genetic diversity between the PCNSL and DLBCL cancer genomes, the existing molecular subtyping methods for PCNSL^14^ and DLBCL^9–12,55,56^ may not be applicable to Chinese PCNSL patients. We assessed the prognostic value of these published molecular subtypes for overall survival (OS) and progression-free survival (PFS) in our Chinese PCNSL cohort. Actionable PCNSL classification can perfectly differentiate prognoses, thereby improving precision medicine strategies. According to cell-of-origin (COO) molecular subtyping^55,56^, there were no significant differences in OS (*P* = 0.42) or PFS (*P* = 0.67) between the germinal center type (GCB) and non-GCB subtypes (**Figure S6A**). Similarly, according to LymphPlex molecular subtyping^12^, no significant differences in OS (P = 0.68) or PFS (P = 0.71) were observed between the BN2-like, MCD-like, EZB-like, N1-like, TP53, and other subtypes (**Figure S6B**). We further categorized our data on the basis of the DLBCL subtyping (C0-C5) proposed by *M. A. Shipp* et al.^10^ Within our cohort, the C4 subtype was associated with shorter OS (*P* = 0.009, P = 0.013) and PFS (*P* = 0.096, P = 0.047) than the C0 and C5 subtypes, respectively (**Figure S6C**). However, the overall efficacy of this DLBCL subtyping system in predicting outcomes was less than satisfactory (OS, P = 0.074). The data were also analyzed via the five subtypes (BN2, EZB, MCD, N1, and others) ^9^ and seven subtypes (A53, BN2, EZB, MCD, N1, ST2, and others)^11^ of DLBCL proposed by *Louis M Staudt* et al. There were no significant differences in OS (*P* > 0.05) or PFS (*P* > 0.05) between BN2, N1, and other categories within the 5-subtype model (**Figure S6D**). Overall, the seven-subtype model (**Figure S6E**) had inferior performance, with no significant differences observed in OS (*P* = 0.61) or PFS (*P* = 0.47). With respect to the four subtypes (CS1-4) of PCNSL proposed by A Alentorn ^14^, our data revealed that the OS (*P* = 0.12) and PFS (*P* = 0.54) of patients with the CS3 subtype were shorter than those of patients with the other three subtypes (**Figure S6F**), but the differences were not significant. Furthermore, the double-hit gene expression signature defines a distinct subgroup of germinal center B-cell-like DLBCL, which is associated with a poor prognosis^13^. However, in this study, there were no significant differences in OS (P > 0.05) or PFS (P > 0.05) between the double- or triple-hit PCNSL patients and the other PCNSL patients, as shown in **Figure S7** (A: all; B: discovery cohort; C: validation cohort). Taken together, these results suggest that the molecular subtyping of PCNSL and DLBCL, as previously published, is not applicable to Chinese PCNSL patients.

### PCNSL molecular subtypes with clinical outcome implications

To establish a molecular classification system for PCNSL, we initially carried out consensus clustering on 58 patients (discovery cohort) who received immunochemotherapy (HD-MTX-based chemotherapy regimen followed by consolidation therapy) via WES data to generate cluster assignments (refer to Methods - Classification model to identify molecular types). WES data from 82 patients (validation cohort) who received the same treatment regimen were used to confirm the molecular clusters. Additionally, WES/WGS sequencing data from 36 newly diagnosed PCNSL patients at Fujian Cancer Hospital and The First Affiliated Hospital of Fujian Medical University were included as an independent external Chinese PCNSL cohort to further validate the molecular clusters^17^.

The molecular classification scheme, presented in **Figure 4A**, **Table S3** and **Figure S8**, was determined independently of clinical information and finalized before the analysis of clinical data, allowing us to analyze the relationships between genetic subtypes and survival in this entire cohort. Finally, EP300, BTG1, MYD88, and IRF4 were included in the molecular subtyping of Chinese PCNSLs.

**Figure 4.**
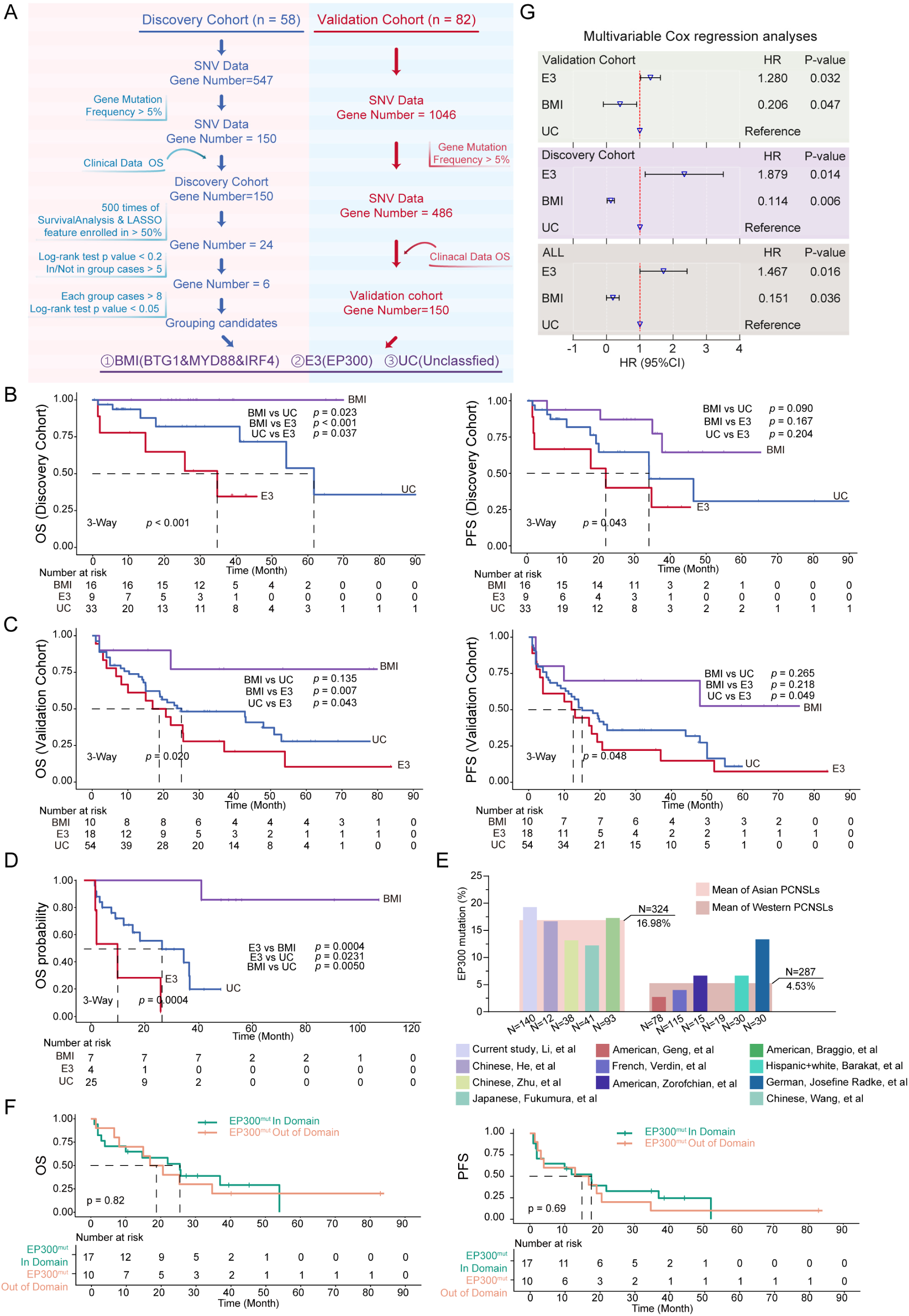
Integrated genetic drivers reveal PCNSL molecular subtypes with clinical outcome implications. A: Schematic of the typing strategies used to identify PCNSL molecular subtypes in the discovery cohort (n = 58) and in the validation cohort (n = 82). B: Kaplan‒Meier estimates of overall survival (OS) and progression-free survival (PFS) among patients belonging to each molecular subtype in the discovery cohort. C: Kaplan‒Meier estimates of overall survival (OS) and progression-free survival (PFS) among patients belonging to each molecular subtype in the validation cohort. D: Kaplan‒Meier estimates of overall survival (OS) among patients belonging to each molecular subtype in an independent external cohort (n=36). E: The prevalence of EP300 mutations in PCNSL patients of different races. F: Kaplan‒Meier analyses for comparisons of the overall survival (OS) and progression-free survival (PFS) of patients with mutations within and those with mutations outside the EP300 domain. G: Hazard ratio estimates of overall survival in PCNSL patients (n = 140).

All nonsynonymous mutations in EP300, BTG1, MYD88, and IRF4 were visualized within the functional domains of the encoded proteins (**Figure S9**). As previously reported^10,57,58^, MYD88 (L265P) was the most prevalent mutation, while EP300, BTG1, and IRF4 all harbored scattered mutations. We evaluated the prognostic significance of these four identified genes for OS and PFS. The survival outcomes of patients with EP300 mutations were significantly poorer than those of patients with wild-type EP300, and patients harboring mutations in either BTG1 (OS, P = 0.017; PFS, P = 0.12), MYD88 (OS, P = 0.016; PFS, P = 0.069), or IRF4 (OS, P = 0.045; PFS, P = 0.018) had favorable OS (**Figure S10A**) and PFS (**Figure S10B**) relative to individuals with their wild-type counterparts.

In the discovery cohort, we identified three subtypes of PCNSL: BMI (n = 16, 27.59%), E3 (n = 9, 15.52%), and UC (n = 33, 56.90%). The E3 subtype was defined as those with mutations in the EP300 gene. The PCNSLs that carried mutations in at least two of the three genes, BTG1, MYD88, and IRF4, were characterized as the BMI subtype. The other PCNSLs that did not fit into either of these categories were classified under the UC subtype (Unclassified). The three subtypes differed significantly in OS (*P* < 0.001) and PFS (*P* = 0.043), the BMI subtype (*P* < 0.001, *P* = 0.023) had much more favorable outcomes than the E3 and UC subtypes did, and the E3 subtype (*P* < 0.001, *P* = 0.037) had far worse outcomes than the BMI and UC subtypes did (**Figure 4B**). These differences between the three subtypes of PCNSL were detected in the validation cohort (OS, *P* = 0.020; PFS, *P* = 0.048; n = 82, tumor-only sample), and patients with E3 had significantly shorter survival times than those with BMI (OS, *P* = 0.007) or UC (OS, *P* = 0.043) (**Figure 4C**). Similar results were observed in the entire cohort (n = 140, OS, *P* < 0.0001; PFS, *P* = 0.0004; **Figure S10C**): 26 patients were classified as BMI (18.57%), 27 as E3 (19.29%), and 87 as UC (62.14%). Furthermore, the raw WES or WGS data, along with follow-up data from an independent cohort of 36 Chinese patients with PCNSL^17^, were obtained. The three identified subtypes significantly differed in OS (P = 0.0004; **Figure 4D**). Specifically, the BMI subgroup (P = 0.0004, P = 0.005) had notably more favorable outcomes than the E3 and UC subgroups did. Conversely, the E3 subtype (P = 0.0004, P = 0.0231) had considerably worse outcomes than either the BMI or UC subgroup did. These results further validate the robustness of our molecular classification.

BTK inhibitors have been shown to improve the prognosis of PCNSL patients by blocking B-cell receptor signaling pathways, which influence the growth and survival of B cells. Consequently, we further analyzed whether the use of BTK inhibitors affects the robustness of our classification. There were no significant differences in OS (P > 0.05, **Figure S11A**) or PFS (P > 0.05, **Figure S11B**) between patients who received BTK inhibitors (n = 8) and those who did not receive BTK inhibitors (n = 132). Additionally, after initial treatment options (HD-MTX combined with idarubicin (IDA), HD-MTX combined with rituximab (R), HD-MTX combined with IDA and R, or a combination of BTK inhibitors) and consolidated therapy (WBRT, stem cell transplant) were adjusted, multivariable Cox regression analysis revealed that the use of different treatment options did not affect the robustness of our classification (**Table S4**).

For the E3 subtype, the mean mutation prevalence in the PCNSLs of Asian patients^3,16,17^ (16.98%) was significantly higher than that in the PCNSLs of Western patients^4,5,14,59–61^ (4.53%) (**Figure 4E**). Patients with the E3 subtype had significantly poorer survival, which was not affected by the mutation site (OS, P = 0.82; PFS, P = 0.69; **Figure 4F**). Univariate Cox regression analysis revealed that PCNSL molecular subtypes are associated with clinical outcomes (**Table S5**), which was also confirmed by multivariate Cox regression hazard ratio analysis after adjusting for important confounders (age, IELSG, MSKCC, LDH levels, deep brain location, and CSF protein levels) (**Figure 4G)**. These findings further confirmed that PCNSL molecular subtypes are associated with clinical outcomes. The key demographic and clinical characteristics of the three MS patients are summarized in **Table S6**.

We also constructed a Sankey diagram to visualize the correspondence between our samples and the previously described genetic subtypes of PCNSL and DLBCL (**Figure S12**). The Chinese PCNSL subtypes were significantly distinct from those previously described. Taken together, these results indicate that three unique molecular subtypes of Chinese PCNSL are associated with clinical outcomes.

### Genomic landscape and tumor microenvironment across PCNSL subtypes

We further aimed to determine whether these three molecular subtypes of Chinese PCNSL patients could affect the mutational landscape, oncogenic pathways and tumor microenvironment. Intergroup mutation distribution analysis was performed to identify unique and shared mutations. A total of 6,102 (median variants per sample = 196.5), 41,548 (median variants per sample = 1719), and 73,440 mutated genes (median variants per sample = 918) were identified in the BMI, E3, and UC subtypes, respectively, with 518 mutated genes shared among the three subtypes (**Figures 5A** and **S13A**). Key candidate cancer genes in the 140 PCNSL patients, grouped by these three subtypes, are presented in **Figure 5B**. The mutation frequencies of EP300, MYD88, IRF4, FAT4, KMT2C, KMT2A, MPEG1, PRKDC, SPEN, CARD11, RANBP2, NUMA1, NSD1, PASK, USP6, PTPN13, SLX4, ADGRA2, TET1, and NOTCH2 were significantly different (P< 0.05) among these three subtypes. Arm-level CNAs among the samples of the three subtypes are displayed in **Figure 5C**, showing significant differences in recurrent deletions (in 6q and 12p) but not in amplifications (**Figure 5D**). The results of significant recurrent focal amplifications and deletions were visualized by GISTIC 2.0 in the BMI, E3, and UC subtypes (**Figure S13B**). There was no significant difference (*P* > 0.05) in the MATH (**Figure 5E**) or MSI (**Figure 5F**) scores among the three subtypes, but the TMB was greater in the E3 subgroup than in the BMI (*P* < 0.0001) or UC subgroup (*P* < 0.001) (**Figure 5G**). Furthermore, the VAF was lower in the E3 subgroup than in the BMI (*P* < 0.0001) or UC subgroup (*P* < 0.001) (**Figure 5H**).

**Figure 5.**
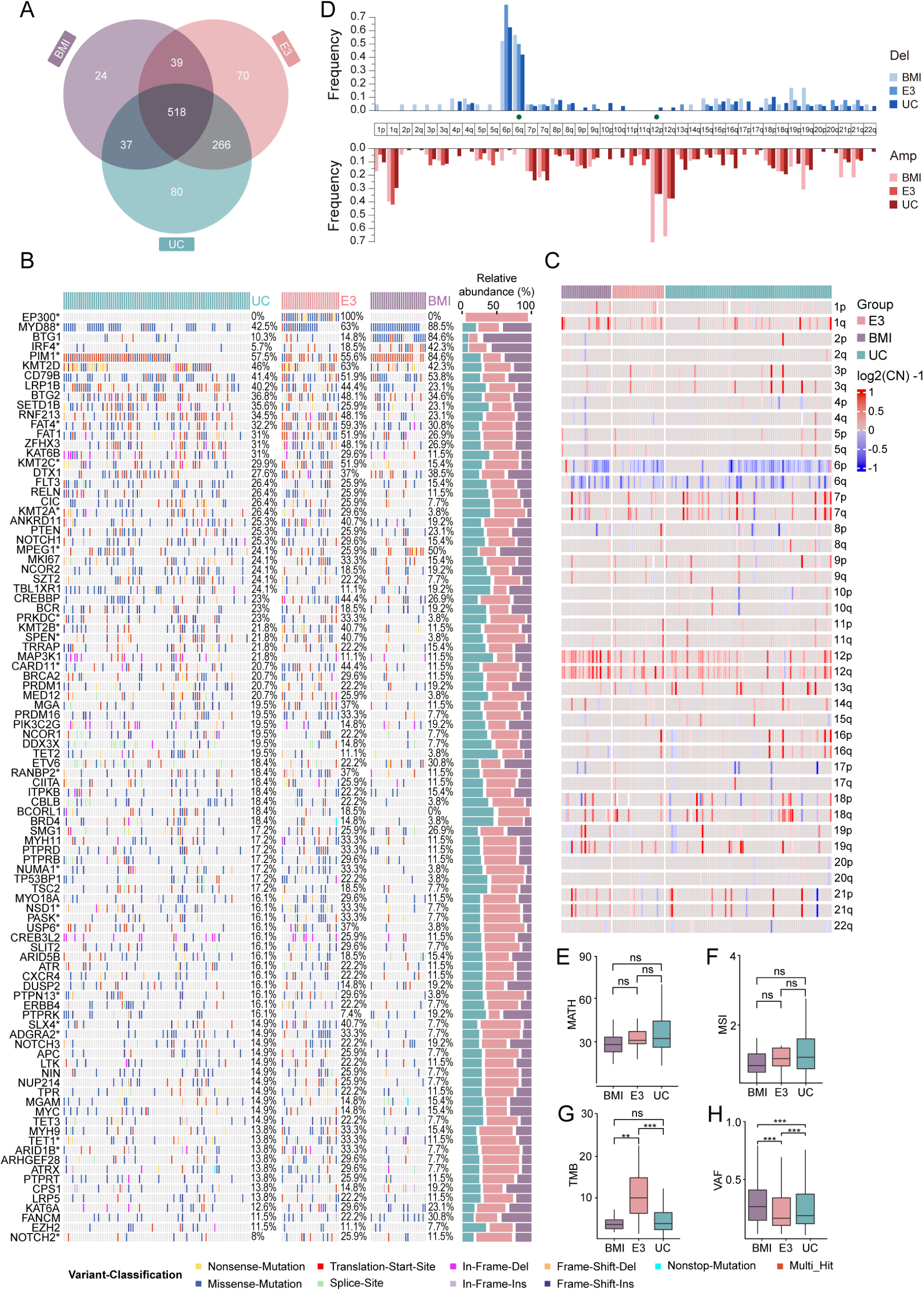
Mutation profiles across three molecular subtypes of PCNSL. A: Venn diagram showing unique and shared mutations among the three identified PCNSL subtypes. B: Number and frequency of recurrent mutations and the gene‒sample matrix of recurrently mutated genes among the three PCNSL subtypes. The relative abundance across the molecular subtypes is displayed on the right. C: Arm-level copy number alterations among the three molecular subtype samples are displayed. D: The frequencies of amplifications and deletions were compared among the three molecular subtype samples. E: The levels of mutant-allele tumor heterogeneity (MATH) were compared among the three molecular subtype samples. F: The levels of microsatellite instability (MSI) were compared among the three MS samples. G: The tumor mutational burden (TMB) was compared among the three MS samples. H: The levels of variant allele frequency (VAF) were compared among the three MS samples. Independent-samples t tests and chi-squared tests were used. \**P* < 0.05; \*\**P* < 0.01; \*\*\**P* < 0.001; ns *P* > 0.05.

The contributions of mutational signatures across the three subtypes are illustrated in **Figure S13C**. The three signatures extracted from the E3 subtype samples displayed cosine similarities of 82.0%, 93.8%, and 95.2% to the COSMIC signatures 6, 10a, and 45, respectively (**Figure S13D**). In contrast, those extracted from BMI subtype samples presented similarities of 77.3%, 81.7%, and 58.3% to COSMIC signatures 5, 84, and 87, respectively (**Figure S13D**). The signatures from the UC subtype samples were 80.8%, 70.3%, and 91.7% and similar to the COSMIC signatures 5, 1, and 45 (**Figure S13D**), respectively.

Hematoxylin–eosin (H&E) and immunohistochemical (IHC) staining of CD2, CD5, CD10, CD19, CD20, CD79a, Ki-67, BCL2, BCL6, MUM1, c-Myc, and p53 from samples of different PCNSL subtypes revealed significant malignant progression in patients with the E3 subtype of PCNSL, as shown in **Figure S14A** and **Table S7**. This is evidenced by the formation of poorly differentiated and aggressively growing tumors (Ki-67), which contrasts with the other two subtypes. Immunostaining revealed a greater presence of CD2+, CD5+, CD10+, CD19+, CD20+, and CD79a+ cells in the E3 subtype. Additionally, a greater presence of BCL2+, BCL6+, MUM1+, and c-Myc+ cells, which are markers associated with the diagnosis of B-cell lymphomas^62^, was observed in the E3 subtype.

To explore potential therapeutic strategies for the genetic subtypes of PCNSL, we examined groups of genetic aberrations that target oncogenic signaling pathways (**Figure S14B**). Genetic events affecting NF-κB regulators that negatively regulate the stability of NF-κB-dependent mRNAs were detected in 85.2% and 84.6% of the E3 and BMI subtype samples but not in the UC subtype samples (P <0.05). These findings suggest that the E3 and BMI subtypes may be more responsive to BTK inhibitors^63^. The PI3 kinase pathway, a pathway that can indirectly activate NF-κB, is genetically altered in 66.7% of E3 patients^64^. We also noted a higher frequency of genetic alterations in chromatin histone modifiers (100%), RTK signaling (77.8%), protein homeostasis/ubiquitination (66.7%), cell cycle pathways (77.8%), genome integrity (82.5%), Wnt/B-catenin signaling (59.3%), the chromatin SWI/SNF complex (70.4%), and splicing (37%) in E3 patients. Therefore, a combination of cyclin D-Cdk4,6 and PI3 kinase inhibitors might be beneficial for patients with the E3 subtype.

Taken together, these results suggest that the three molecular subtypes of PCNSL in Chinese patients each have a unique genomic landscape, tumor microenvironment, and oncogenic pathways.

### Mutational landscape and oncogenic pathways in PCNSLs with different sites of onset

Next, we aimed to determine whether the site of PCNSL onset in Chinese patients influences the mutational landscape and oncogenic pathways. **Figure 6A** shows that the areas most often affected by PCNSL were the basal ganglia, brainstem, cerebellum, corpus callosum, frontal lobe, occipital lobe, parietal lobe, temporal lobe, thalamus, and ventricles, as well as combinations of multiple sites. The most frequently involved sites of PCNSL were the frontal lobe (22.14%), temporal lobe (15.71%), and basal ganglia (8.57%).

**Figure 6.**
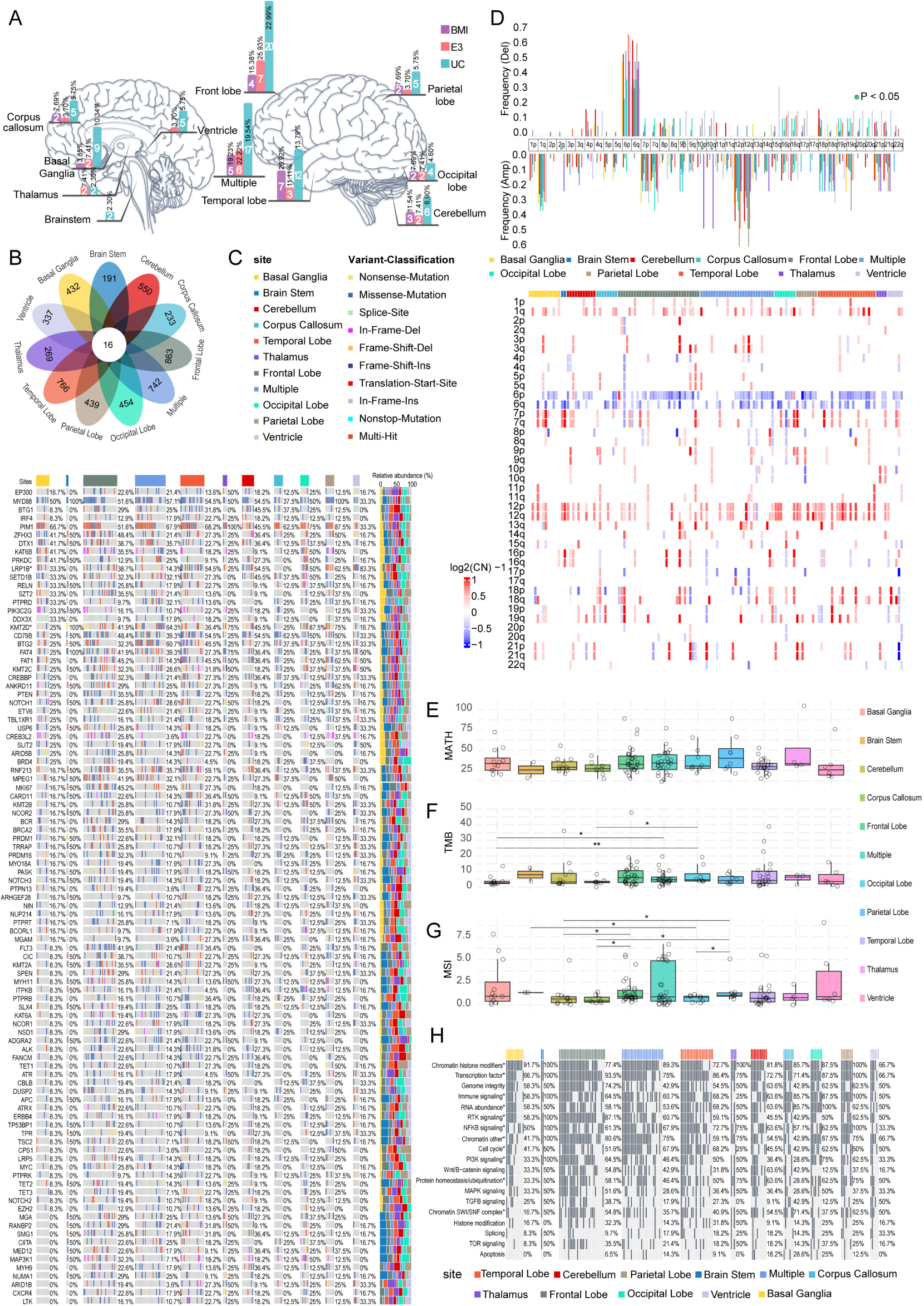
Mutational profiles across different sites of onset in PCNSL. A: The number and percentage of PCNSL patients with various molecular subtypes at different disease onset sites. B: Venn diagram of unique and shared mutations among PCNSLs with different sites of onset. C: The number and frequency of recurrent mutations, along with a gene‒sample matrix of recurrently mutated genes in PCNSL patients at different sites of onset, are presented. The relative abundance across the molecular subtypes is displayed on the right. D: Arm-level copy number alterations across different sites of onset are displayed. The frequencies of amplifications and deletions were compared. E: The levels of mutant-allele tumor heterogeneity (MATH) were compared among the samples representing different sites of onset. F: Comparisons of the tumor mutational burden (TMB) among the different sites of onset samples were performed. G: The levels of microsatellite instability (MSI) were compared among the samples representing different sites of onset. H: Pathways affected by oncogenes in PCNSL patients with different sites of onset. The chi-squared test and one-way ANOVA were used. \**P* < 0.05.

First, we found that there was no significant difference in the proportion of different disease sites among the three subtypes (chi-square test, corpus callosum, P = 0.887; basal ganglia, P=0.645; thalamus, P= 0.317; brainstem, P= 1.000; ventricle, P= 0.631; front lobe, P= 0.576; multiple, P= 0.949; temporal lobe, P= 0.214; cerebellum, P= 0.831; occipital lobe, P= 0.778; parietal lobe, P= 0.887). BMI-related tumors were slightly more prevalent in the temporal lobe (7 out of 26 patients vs. 15 out of 114 patients, chi-square test P = 0.082) than other subtypes were. Conversely, E3 events were likely to be more frequent in the thalamus (2 out of 27 cases vs. 2 out of 113 cases, chi-square test P = 0.113). Larger studies are needed to confirm this observation.

Next, we explored whether the tumor sites differed between the discovery and validation cohorts. As shown in **Figure S15A**, there was no significant difference in the proportions of different disease sites between the discovery and validation cohorts (chi-square test P = 0.610). Furthermore, the proportions of deep brain lesions in the discovery and validation cohorts were not significantly different (P = 0.478).

Sixteen mutated genes were represented among all the sites (**Figure 6B**). Key candidate cancer genes in the 140 PCNSL patients, grouped by site of disease onset, are presented in **Figure 6C**. Mutations in the PIM1, CD79B, and MYD88 genes were relatively common across all PCNSL onset sites. The frontal lobe had the highest mutation frequencies of five genes: PIM1 (51.61%), MYD88 (51.61%), and ZFHX3 (48.39%). In the temporal lobe, the most prevalent mutations were in PIM1 (68.18%), SC1. In the basal ganglia, the genes with the highest mutation frequencies were PIM1 (66.67%), MYD88 (50%), ZFHX3 (41.67%), and DTX1 (41.67%). The mutation frequencies of LRP1B and KMT2D were significantly different (P< 0.05) among these sites. Arm-level CNAs among the samples of these sites are displayed in **Figure 6D**, and significant differences in both recurrent amplifications (2q, 10q, and 10p) and deletions (9p) were observed. The difference in the MATH score was significant (*P* < 0.05) (**Figure 6E**) across these sites, but there was no significant difference (*P* > 0.05) in the TMB (**Figure 6F**) or MSI (**Figure 6G**) score between several sites.

Next, we further divided the tumor sites into deep brain and shallow brain tumor subgroups. A total of 854 (81%) mutated genes were represented between the deep brain and shallow brain tumor subgroups (**Figure S15B**). The mutational landscape between deep brain and shallow brain tumors is shown in **Figure S15C**. The mutation frequencies of LRP1B, FAT1, KMT2B, CIC, PTPRB, MAP3KI, ARID1B, and CPS1 in the deep brain and shallow brain tumors were significantly different (P< 0.05). Arm-level CNAs between deep brain and shallow brain tumors are displayed in **Figure S15D**, and no significant differences (P >0.05) in either recurrent amplification or deletion were observed. There was no significant difference (*P* > 0.05) in the MATH (**Figure S15E**), TMB (**Figure S15F**), or MSI (**Figure S15G**) score between deep brain and shallow brain tumors.

We further conducted pathway enrichment analysis (**Figure 6H**) and revealed significant involvement of NF-κB signaling in the tumors of patients with onset in the parietal lobe and brainstem, each with an involvement rate of 100%. We also noted a significant difference (P <0.05) in the frequency of genetic alterations in transcription factors, other chromatin, immune signaling, NF-κB signaling, chromatin histone modifiers, RTK signaling, PI3K signaling, RNA abundance, the chromatin SWI/SNF complex, protein homeostasis/ubiquitination, and the cell cycle among these sites. Furthermore, we noted a significant difference (P <0.05) in the frequency of genetic alterations in the chromatin SWI/SNF complex but not in the other pathways (**Figure S15H**). Taken together, these results indicate that the mutational landscape in Chinese patients with PCNSL varies between sites of onset.

## Discussion

Owing to the genetic, phenotypic, and tumor microenvironment heterogeneity of PCNSL, identifying classification and prognostic biomarkers for patients is extremely challenging, which in turn makes developing effective new therapies challenging. Here, we performed a study of 176 Chinese PCNSL patients that was adequately powered to expand the genomic landscape and explore its clinical significance utilizing an unprecedented sample size and a multicenter approach. We defined recurrent mutations, CNAs, and associated cancer candidate genes in Chinese PCNSL patients and compared these comprehensive genetic signatures with those of PCNSL patients and systemic DLBCL patients of other races, revealing that genetic heterogeneity is a defining feature of PCNSL. Our results highlight the complexity of PCNSLs, which have a median of 19.06 mutations/Mb and a median of 949.5 variants per sample.

Although the genetic variation profiles of the PCNSL discovery and validation samples were similar, notable differences were evident in the mutation frequencies of certain genes (e.g., NOTCH1, NOTCH2, and KMT2C), the TMB, the MSI, and the MATH score. These discrepancies might stem from variations in specimen types (fresh tissue versus FFPE) and the absence of paired samples in the validation cohort^65^. Furthermore, genetic heterogeneity between patient populations^15^ and biological differences between PCNSL and DLBCL^13^ may be the main reasons why the published classifications for DLBCL and PCNSL are not applicable to Chinese PCNSL patients. Currently, the classification frameworks for systemic DLBCL offer valuable insights into the underlying biology based on molecular mechanisms. However, it is important to note that biological insights do not necessarily correlate with clinical outcomes, especially given that treatment paradigms may evolve over time.

More importantly, in our study, we identified three robust molecular subtypes of PCNSL in Chinese patients: BMI, E3, and UC. Distinct clinical outcomes, activated cellular pathways, and alterations in the genetic landscape distinguish the three subtypes and could be utilized to personalize therapy for patients with different subtypes. For the E3 subtype, the mean mutation rate was significantly higher in the PCNSLs of Chinese and Japanese patients (16.88%) than in those of Western PCNSL patients (4.53%). This difference may be attributed to variations in age, lifestyle, and EBV infection rates^17,41,66^. Patients with the E3 subtype had significantly poorer survival outcomes, a finding that is consistent with reported findings in DLBCL^67^. EP300 and CREBBP are two closely related members of the KAT3 family of histone acetyltransferases that function similarly^68^. However, in this study, the survival patients with EP300 mutations was significantly poorer, whereas CREBBP mutations did not contribute to an inferior prognosis (**Figure S4**). This discrepancy might stem from the distinct roles of EP300 and CREBBP in PCNSL. Specifically, in a recent study, EP300, but not CREBBP, was reported to play an essential role in supporting the viability of classical Hodgkin’s lymphoma by directly modulating the expression of the oncogenic MYC/IRF4 network, surface receptor CD30, immunoregulatory cytokine interleukin 10, and immune checkpoint protein PD-L1^69^. The molecular mechanisms through which EP300 mutations facilitate the progression of PCNSL are still not understood. We plan to explore these mechanisms in a forthcoming study. PCNSLs that carried mutations in at least two of three genes, namely, BTG1, MYD88, and IRF4, were characterized as the BMI subtype with favorable survival outcomes. The prognostic significance of MYD88 mutations in PCNSL patients remains inconclusive. While several studies have reported no effect on OS^70–73^, only two have reported an unfavorable outcome^74,75^. In line with our findings, *Olimpia* et al.^76^ and *Claudio* et al.^77^ have indicated that the survival time is significantly longer in patients with PCNSLs harboring MYD88 mutations. This discrepancy may be attributed to heterogeneity in genetics, phenotype, and the tumor microenvironment across various patient populations; variances in sample size; and the adjustment of confounding variables, including age and treatment methods. In this large study, we found that Chinese PCNSL patients harboring MYD88 mutations have longer survival times. However, further research is needed to verify these findings in diverse populations. The associations of IRF4 and BTG1 mutations with clinical outcomes in PCNSL patients have not been previously reported. This results of this study revealed that mutations in both IRF4 and BTG1 were associated with favorable outcomes. IRF4 has been identified as a driver oncogene that transcriptionally regulates downstream target genes, such as MYC, and coordinates the transcriptional program with NF-κB references^78–81^. Numerous in vitro and clinical studies have highlighted the abnormal overexpression and oncogenic roles of IRF4 in various mature lymphoid neoplasms^81–84^. Therefore, we hypothesized that the reduced expression and/or decreased transcriptional activity of IRF4 resulting from such mutations may explain the association of IRF4 mutations with favorable survival outcomes in PCNSL patients. BTG1 mutation has been associated with extranodal dissemination and unfavorable outcomes in B-cell lymphoma patients^85–87^. However, in this study, BTG1 mutation was linked to favorable outcomes in PCNSL patients. In a substantial discovery cohort consisting of 533 patients with B-cell precursor acute lymphoblastic leukemia, *Blanca* and colleagues^88^ reported that deletions of BTG1 alone did not affect prognosis. The complete remission rate among acute myeloid leukemia patients with low BTG1 expression was significantly greater than that among patients with high BTG1 expression^89^. The molecular mechanisms through which mutations in BTG1 decelerate the progression of PCNSL warrant further research.

This study has several limitations. First, the proportion of unclassified PCNSLs (UC subtypes) is high, possibly due to the lack of multiomics data needed for classification. However, the three-class classification based on WES proposed in this study has strong clinical practicality and translational value, as it relies solely on the four genes identified through WES. In other words, clinicians can predict patient outcomes and further perform individualized management of PCNSLs by detecting mutations in these four genes without the need for additional multiomics testing. Second, the three identified molecular subtypes can be used to predict patient outcomes. However, it remains challenging to determine whether these three molecular subtypes can still be used to accurately distinguish biological subtypes without additional RNA-seq data layers.

In summary, by performing genomic sequencing on tumor specimens from 176 Chinese PCNSL patients, we identified three molecular subtypes that can be used to predict patient outcomes. These genetic signatures associated with PCNSL outcomes illuminate the path toward individualized management of PCNSL patients and the discovery of novel treatment strategies for this challenging disease.

## Data Availability

All data produced in the present study are available upon reasonable request to the authors.

## Data and code availability

The raw sequence data have been deposited in the Genome Sequence Archive^90^ in the National Genomics Data Center^91^, China National Center for Bioinformation/Beijing Institute of Genomics, Chinese Academy of Sciences (GSA-Human HRA006122), which is publicly accessible at https://ngdc.cncb.ac.cn/gsa-human. Any additional information required to reanalyze the data reported in this paper is available from the lead contact upon request.

The raw sequencing data of the DLBCL WES data are accessible from the TCGA database and were reanalyzed via pipelines and filter settings that were identical to those used in the present study. The raw WES data of the Japanese PCNSL patients were obtained from the Japanese Genotype–Phenotype Archive (JGA, http://trace.ddbj.nig.ac.jp/jga) and reanalyzed via pipelines and filtering settings that were identical to those used in the present study, which is hosted by DDBJ under the accession number JGAS00000000021^3^. The raw sequencing WES/WGS data of an external independent Chinese PCNSL cohort (n = 36)^17^ were obtained from the China National Center for Bioinformation/Beijing Institute of Genomics, Chinese Academy of Sciences and reanalyzed via pipelines and filtering settings that were identical to those used in the present study, under the accession number GSA-Human HRA002475.

The frequencies of recurrent genetic alterations in DLBCLs^49^ and French PCNSLs^14^ are accessible from the manuscript or supplemental information and were not reanalyzed via identical pipelines and filtering settings as those used in the present study.

## Acknowledgment

This study was funded by the National Natural Science Foundation of China (82302582), the Shanghai Municipal Health Commission Project (20224Y0317), the Shanghai Science and Technology Commission (17430750200), the Join Breakthrough Project for New Frontier Technologies of Shanghai Hospital Development Center (SHDC12016120), and the Youth Medical Talents–Clinical Laboratory Practitioner Program (2022-65).

## Author contributions

SJ. L, CX. L, DH. L, WJ. C, ZG. X, XT. Z, and Y. M conceived and designed the project. SJ. L, DH. L, WJ. C, JZ. C, J. R, and ZG. X collected the clinical samples. SJ. L, JN. W, JZ. C, LY. Z, HW. Y, and Y. S analyzed the WES data and performed bioinformatic analyses. SJ. L, DH. L, J. R, LY. Z, HW. Y, and JN. W integrated the sequencing data, drew the display items. SJ. L, DH. L, and CX. L wrote the manuscript. XT. Z and Y. M oversaw the ethical guidelines and data regulation. WJ. C and Y. M supervised the project. All of the authors contributed to the final version of the paper.

