## Supplementary Figures for "Genomic Landscape and Molecular Subtypes of Primary Central Nervous System Lymphoma"

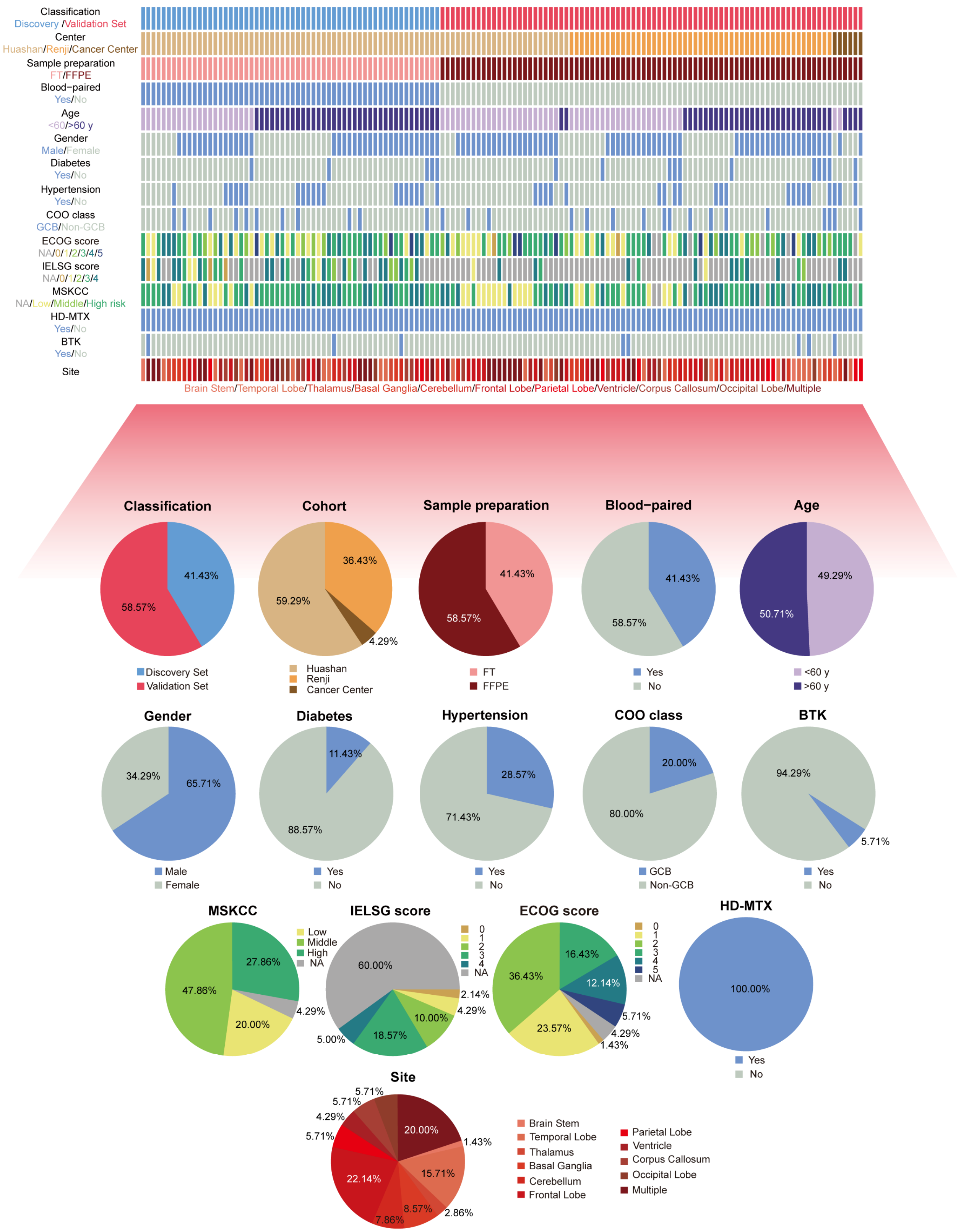


**Figure S1. Basic information of the 140 PCNSL patients**

The key demographic and clinical characteristics of PCNSL patients are summarized in the upper half of the figure, while the proportions of patients with these characteristics are presented in the lower half. Related to Figure 1.

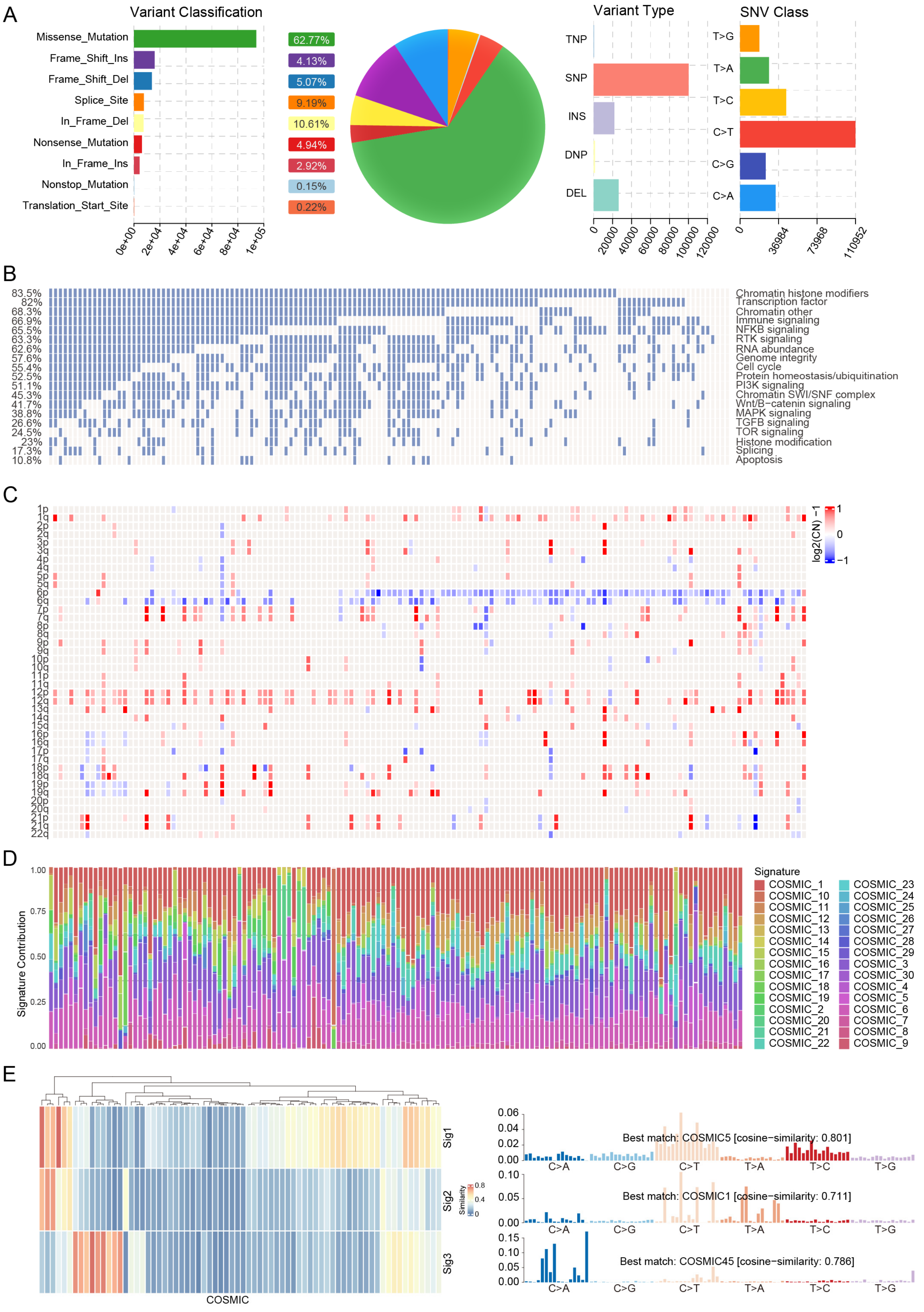
A
Variant Classification
Missense_Mutation
Frame_Shift_Ins
Frame_Shift_Del
Splice_Site
In_Frame_Del
Nonsense_Mutation
In_Frame_Ins
Nonstop_Mutation
Translation_Start_Site
62.77%
4.13%
5.07%
9.19%
10.61%
4.94%
2.92%
0.15%
0.22%
0e+00
2e+04
4e+04
6e+04
8e+04
1e+05
Variant Type
TNP
SNP
INS
DNP
DEL
SNV Class
T>G
T>A
T>C
C>T
C>G
C>A
0
36984
73968
110952
B
Chromatin histone modifiers
Transcription factor
Chromatin other
Immune signaling
NFKB signaling
RTK signaling
RNA abundance
Genome integrity
Cell cycle
Protein homeostasis/ubiquitination
PI3K signaling
Chromatin SWI/SNF complex
Wnt/B−catenin signaling
MAPK signaling
TGFB signaling
TOR signaling
Histone modification
Splicing
Apoptosis
C
log2(CN) −1
D
Signature Contribution
Signature
E
COSMIC
Sig1
Sig2
Sig3
Similarity
Best match: COSMIC5 [cosine−similarity: 0.801]
Best match: COSMIC1 [cosine−similarity: 0.711]
Best match: COSMIC45 [cosine−similarity: 0.786]
C>A
C>G
C>T
T>A
T>C
T>G

**Figure S2. PCNSL mutational landscape and variation signature**

A: Variant classification, type, and single-nucleotide variation class for PCNSL patients (n = 140).

B: Oncogene pathways in 140 PCNSL patients.

C: Arm-level copy number alterations in 140 PCNSL patients.

D: Contribution of mutational signatures in 140 PCNSL patients.

E: The most significant contributions to the PCNSL genome.

Related to Figure 1.

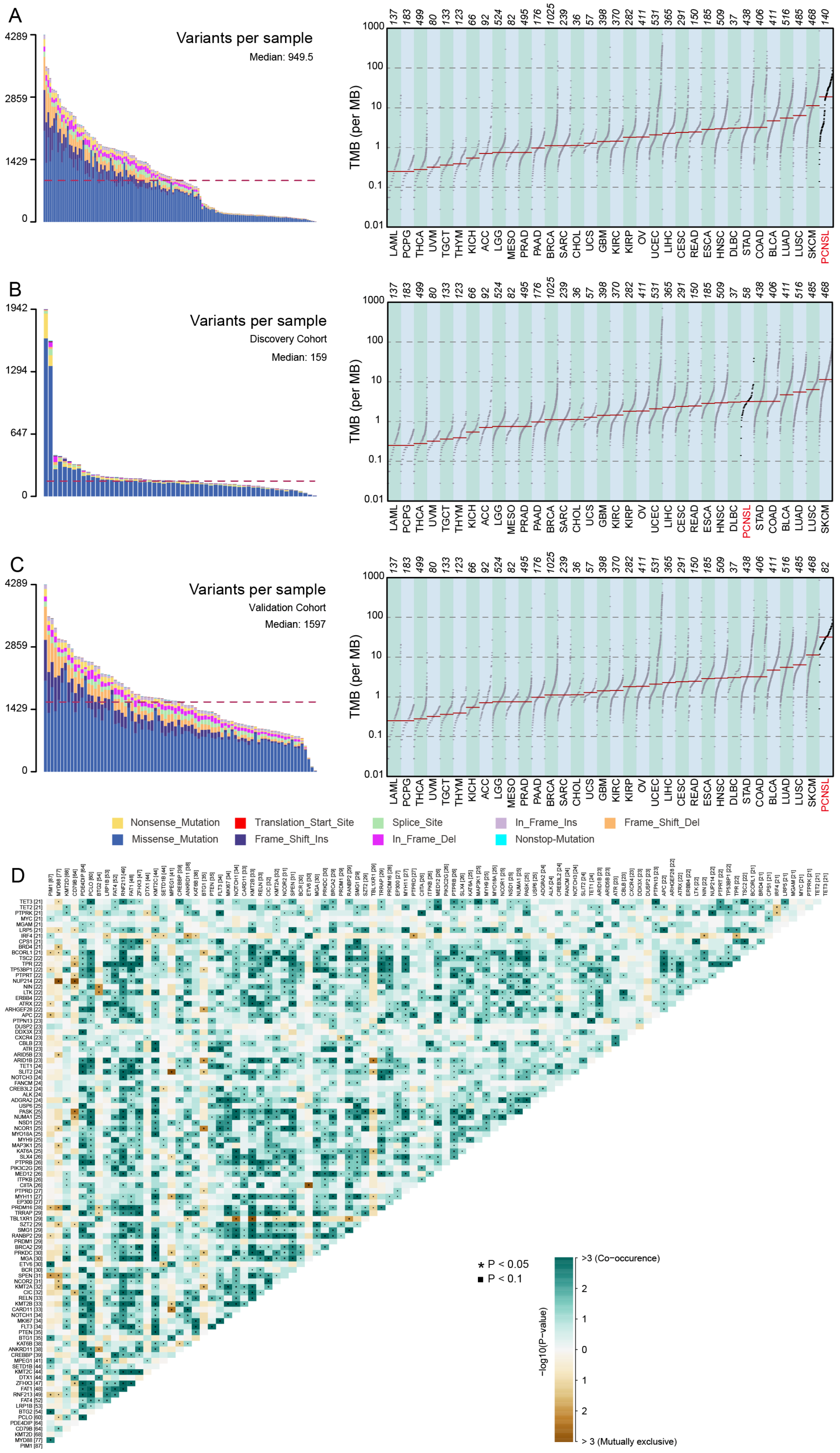
A
Variants per sample
Median: 949.5
4289
2859
1429
0
TMB (per MB)
1000
100
10
1
0.1
0.01
137 183 499 80 133 123 66 92 524 82 495 176 1025 239 36 57 398 370 282 411 531 365 291 150 185 509 37 438 406 411 516 485 468 140
LAML PCPG THCA UVM TGCT THYM KICH ACC LGG MESO PRAD PAAD BRCA SARC CHOL UCS GBM KIRC KIRP OV UCEC LIHC CESC READ ESCA HNSC DLBC STAD COAD BLCA LUAD LUSC SKCM PCNSL
B
Variants per sample
Discovery Cohort
Median: 159
1942
1294
647
0
TMB (per MB)
137 183 499 80 133 123 66 92 524 82 495 176 1025 239 36 57 398 370 282 411 531 365 291 150 185 509 37 58 438 406 411 516 485 468
LAML PCPG THCA UVM TGCT THYM KICH ACC LGG MESO PRAD PAAD BRCA SARC CHOL UCS GBM KIRC KIRP OV UCEC LIHC CESC READ ESCA HNSC DLBC PCNSL STAD COAD BLCA LUAD LUSC SKCM
C
Variants per sample
Validation Cohort
Median: 1597
4289
2859
1429
0
TMB (per MB)
137 183 499 80 133 123 66 92 524 82 495 176 1025 239 36 57 398 370 282 411 531 365 291 150 185 509 37 438 406 411 516 485 468 82
LAML PCPG THCA UVM TGCT THYM KICH ACC LGG MESO PRAD PAAD BRCA SARC CHOL UCS GBM KIRC KIRP OV UCEC LIHC CESC READ ESCA HNSC DLBC STAD COAD BLCA LUAD LUSC SKCM PCNSL
Nonsense_Mutation
Missense_Mutation
Translation_Start_Site
Frame_Shift_Ins
Splice_Site
In_Frame_Del
In_Frame_Ins
Nonstop-Mutation
Frame_Shift_Del
D
* P < 0.05
▪ P < 0.1
>3 (Co−occurence)
2
1
0
1
2
> 3 (Mutually exclusive)
−log10(P−value)

**Figure S3. Genetic variation events and tumor mutational burden (TMB) in the 140 PCNSL samples**

A: Genetic variation events in 140 PCNSL samples and the TMB of PCNSL, ranking 2nd in the TCGA database.

B: Genetic variation events in the discovery cohort of PCNSL patients (n = 58) and the TMB of PCNSL patients, ranking 5th in the TCGA database.

C: Genetic variation events in the validation cohort of PCNSL patients (n = 82) and the TMB of PCNSL patients, ranking 1st in the TCGA database.

D: The mutually exclusive or co-occurrence relationships of these genes.

Related to Figure 1.

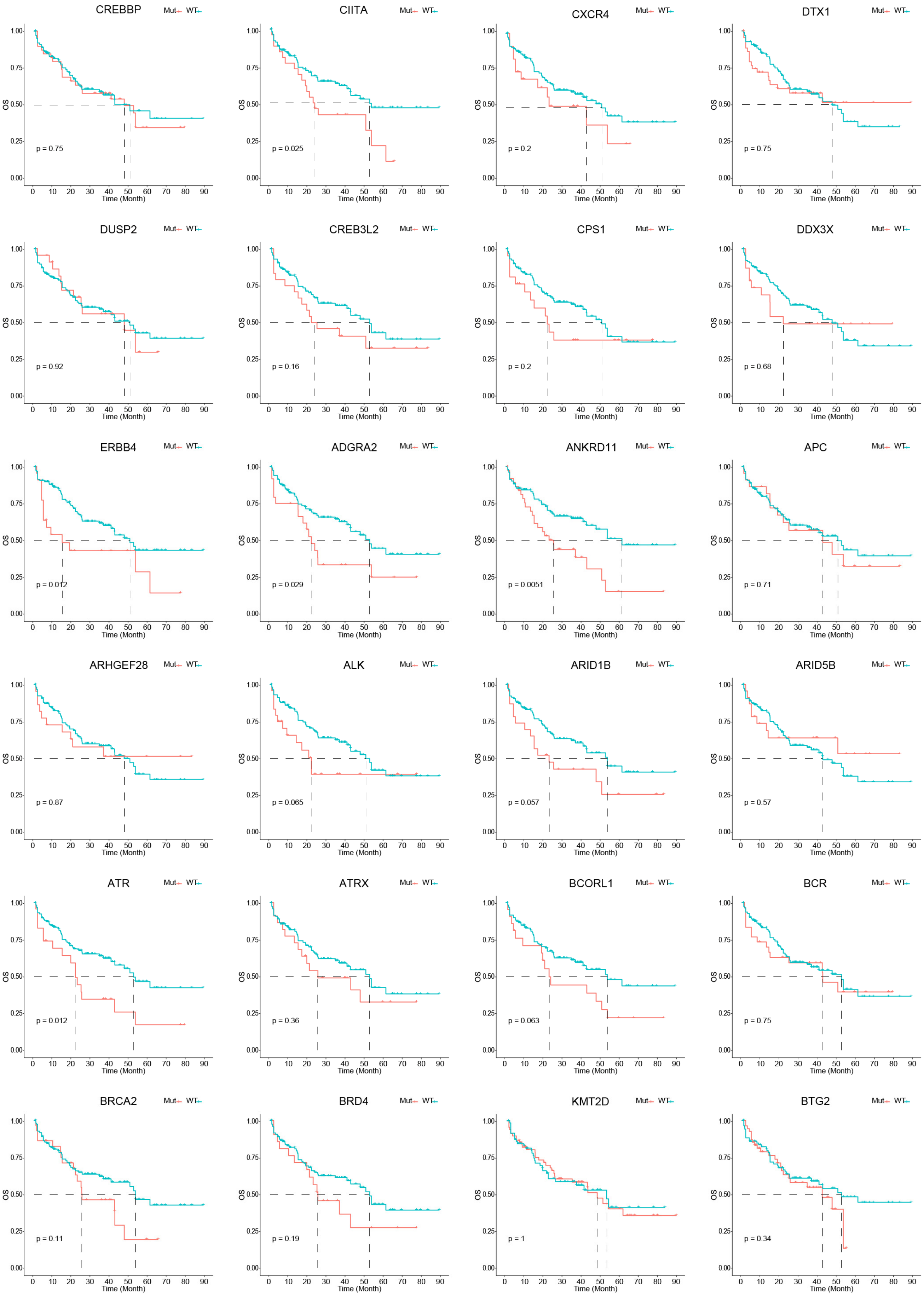

CREBBP
p = 0.75
CIITA
p = 0.025
CXCR4
p = 0.2
DTX1
p = 0.75
DUSP2
p = 0.92
CREB3L2
p = 0.16
CPS1
p = 0.2
DDX3X
p = 0.68
ERBB4
p = 0.012
ADGRA2
p = 0.029
ANKRD11
p = 0.0051
APC
p = 0.71
ARHGEF28
p = 0.87
ALK
p = 0.065
ARID1B
p = 0.057
ARID5B
p = 0.57
ATR
p = 0.012
ATRX
p = 0.36
BCORL1
p = 0.063
BCR
p = 0.75
BRCA2
p = 0.11
BRD4
p = 0.19
KMT2D
p = 1
BTG2
p = 0.34
Mut
WT
OS
Time (Month)

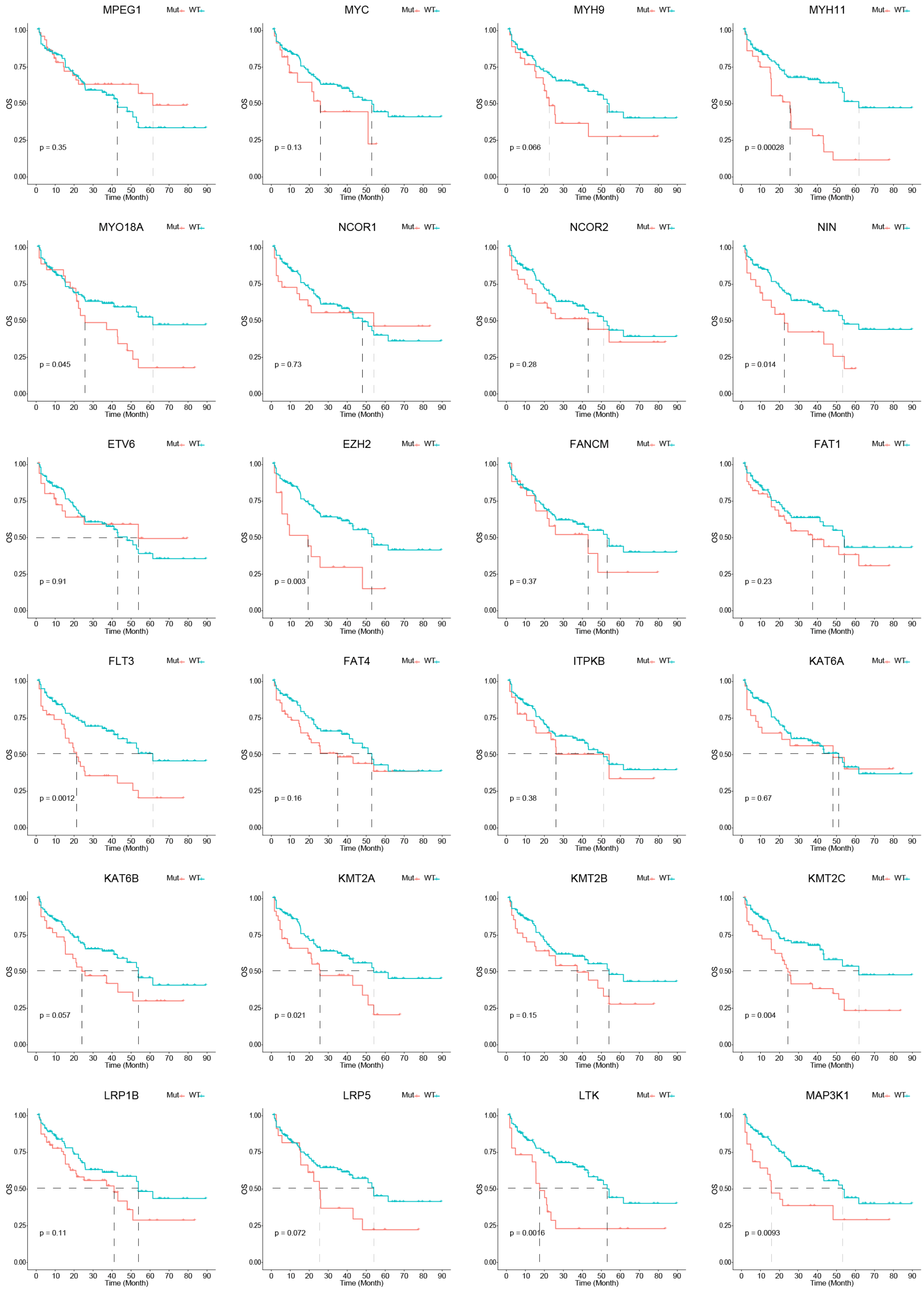
MPEG1
Mut
WT
p = 0.35
MYC
p = 0.13
MYH9
p = 0.066
MYH11
p = 0.00028
MYO18A
p = 0.045
NCOR1
p = 0.73
NCOR2
p = 0.28
NIN
p = 0.014
ETV6
p = 0.91
EZH2
p = 0.003
FANCM
p = 0.37
FAT1
p = 0.23
FLT3
p = 0.0012
FAT4
p = 0.16
ITPKB
p = 0.38
KAT6A
p = 0.67
KAT6B
p = 0.057
KMT2A
p = 0.021
KMT2B
p = 0.15
KMT2C
p = 0.004
LRP1B
p = 0.11
LRP5
p = 0.072
LTK
p = 0.0016
MAP3K1
p = 0.0093
OS
Time (Month)

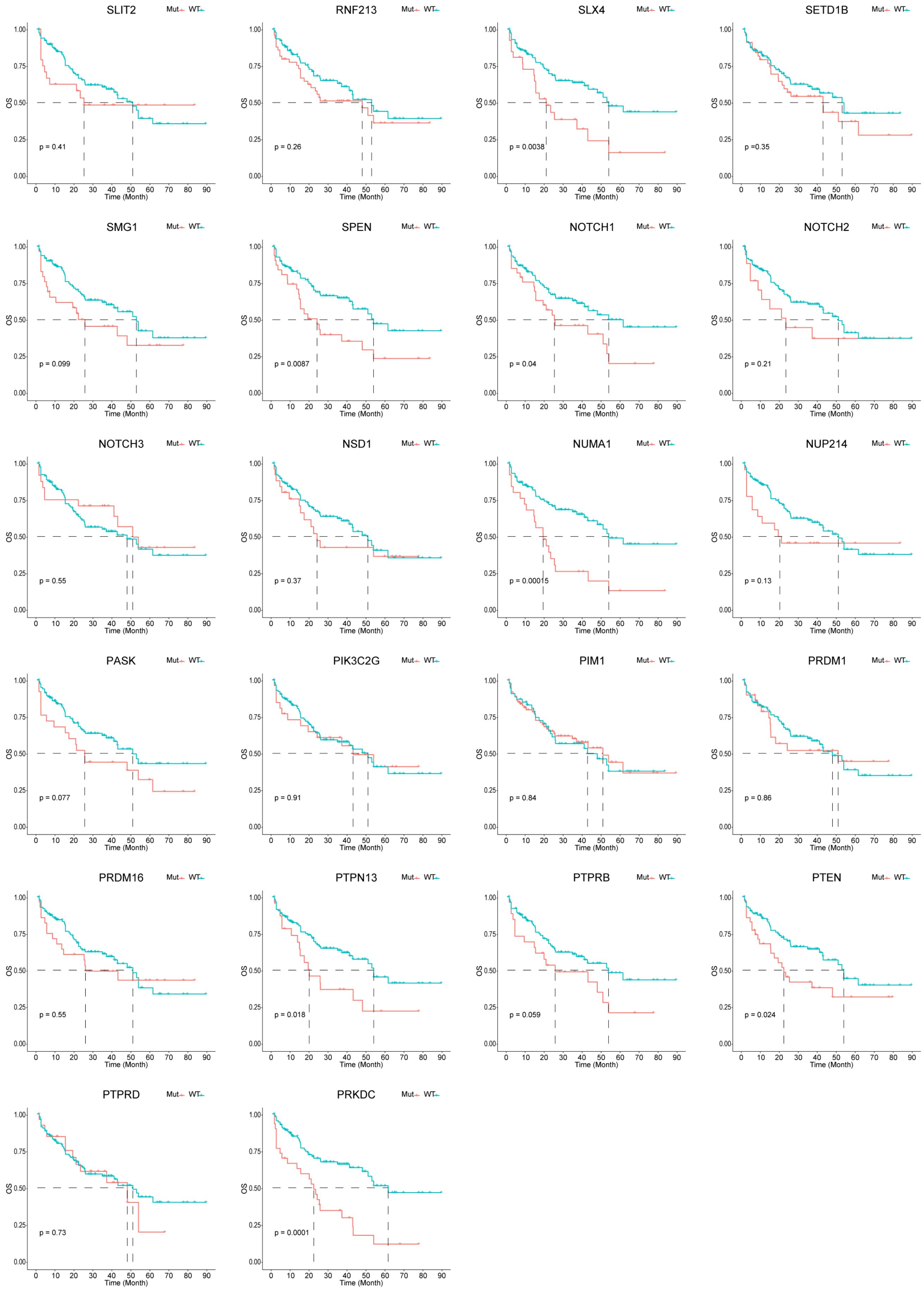
SLIT2
Mut WT
p = 0.41
RNF213
p = 0.26
SLX4
p = 0.0038
SETD1B
p =0.35
SMG1
p = 0.099
SPEN
p = 0.0087
NOTCH1
p = 0.04
NOTCH2
p = 0.21
NOTCH3
p = 0.55
NSD1
p = 0.37
NUMA1
p = 0.00015
NUP214
p = 0.13
PASK
p = 0.077
PIK3C2G
p = 0.91
PIM1
p = 0.84
PRDM1
p = 0.86
PRDM16
p = 0.55
PTPN13
p = 0.018
PTPRB
p = 0.059
PTEN
p = 0.024
PTPRD
p = 0.73
PRKDC
p = 0.0001
OS
1.00
0.75
0.50
0.25
0.00
0 10 20 30 40 50 60 70 80 90
Time (Month)

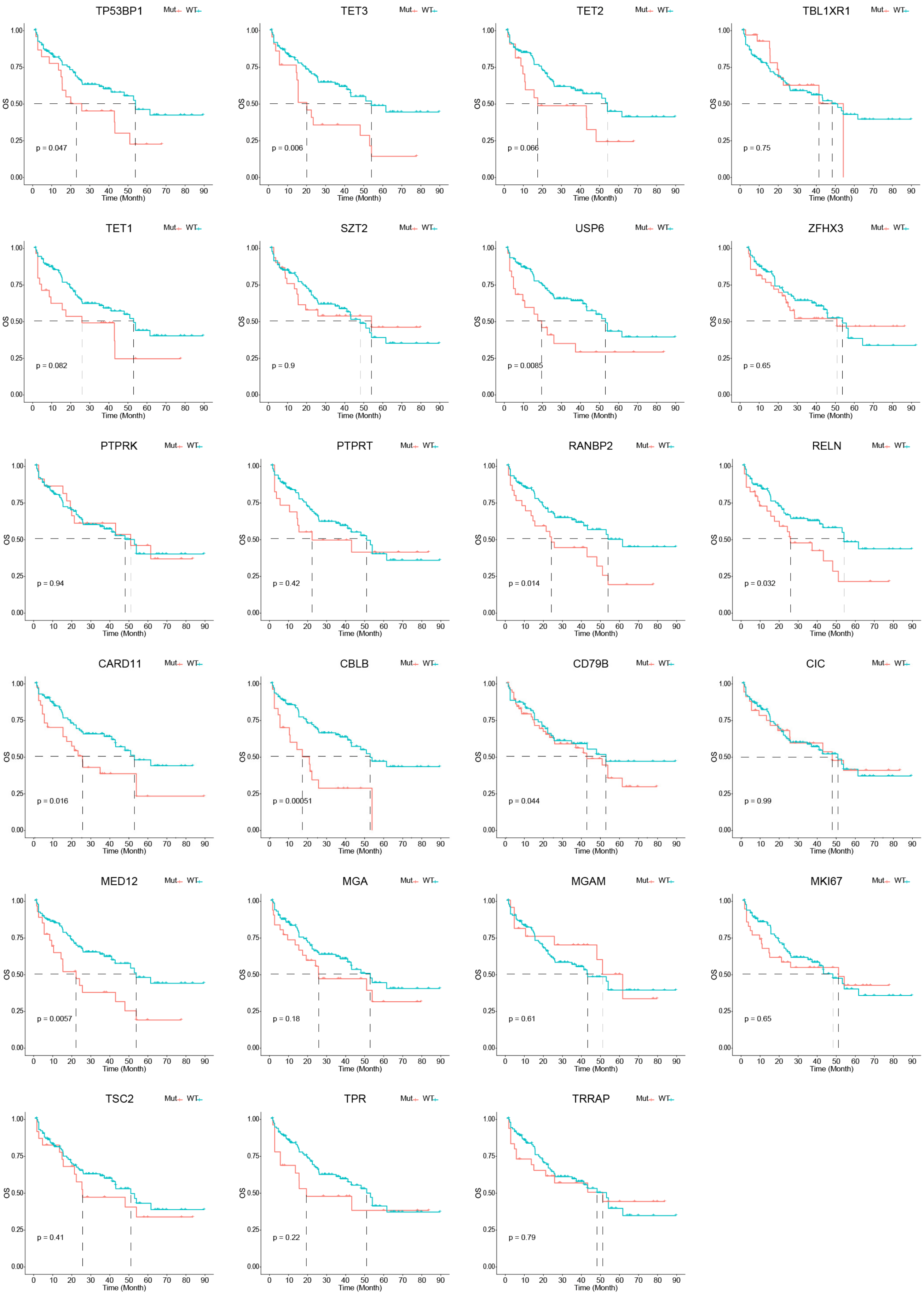

TP53BP1
p = 0.047
TET3
p = 0.006
TET2
p = 0.066
TBL1XR1
p = 0.75
TET1
p = 0.082
SZT2
p = 0.9
USP6
p = 0.0085
ZFHX3
p = 0.65
PTPRK
p = 0.94
PTPRT
p = 0.42
RANBP2
p = 0.014
RELN
p = 0.032
CARD11
p = 0.016
CBLB
p = 0.00051
CD79B
p = 0.044
CIC
p = 0.99
MED12
p = 0.0057
MGA
p = 0.18
MGAM
p = 0.61
MKI67
p = 0.65
TSC2
p = 0.41
TPR
p = 0.22
TRRAP
p = 0.79
Mut
WT
OS
Time (Month)

**Figure S4.** Kaplan–Meier estimates of overall survival associated with candidate cancer genes with mutation frequencies exceeding 15% and the NOTCH2, and EZH2 genes. Related to Figure 1.

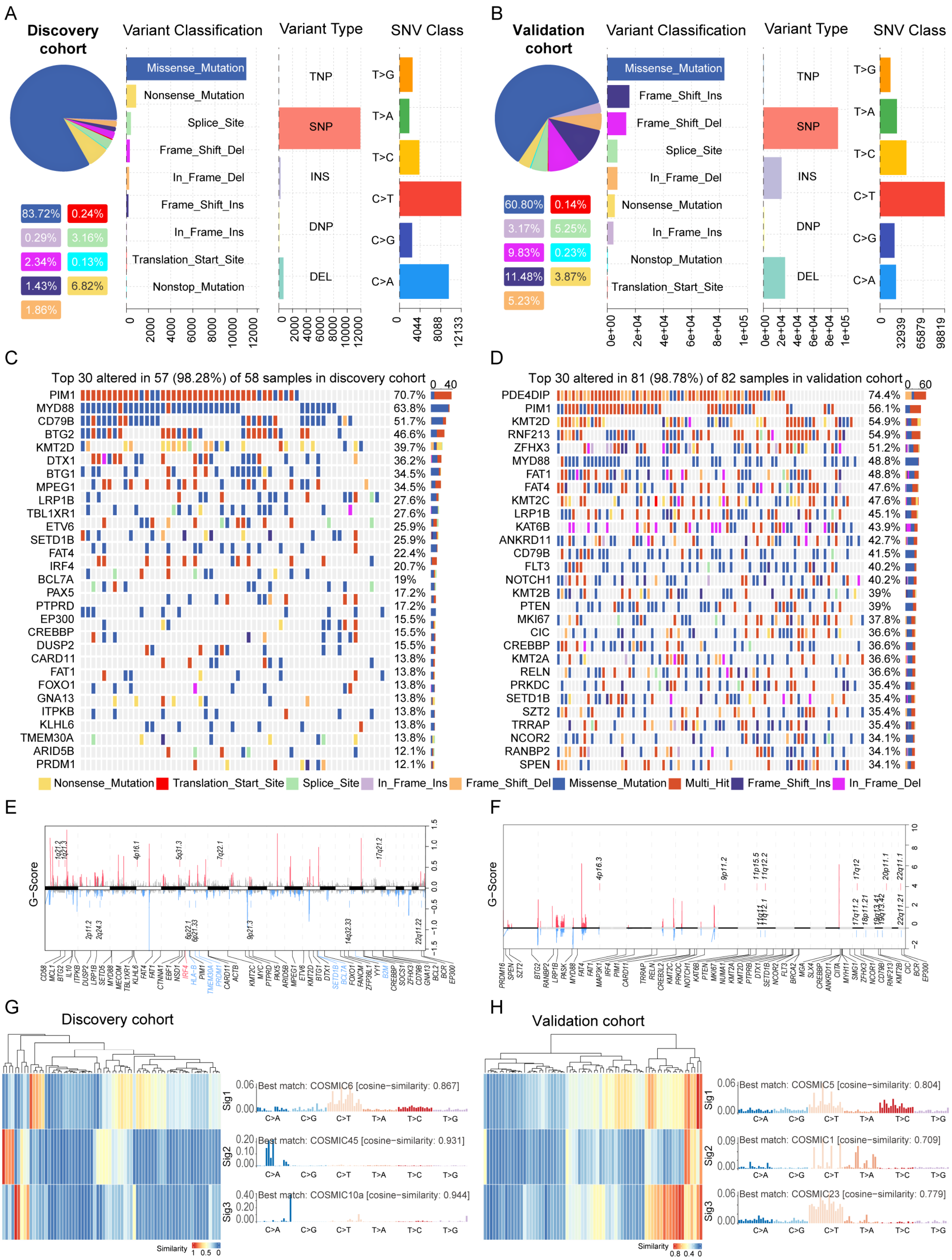

**Figure S5. Mutation Profiles and Signatures of the Discovery and Validation Cohorts**

A: Variant classification, type, and single-nucleotide variation (SNV) class in the discovery cohort of PCNSL patients (n = 58).

B: Variant classification, type, and single-nucleotide variation (SNV) class in the validation cohort of PCNSL patients (n = 82).

C: Number and frequency of recurrent mutations, along with a gene–sample matrix of recurrently mutated genes, ranked by mutation frequency in the discovery cohort of PCNSL patients (n = 58).

D: Number and frequency of recurrent mutations, along with a gene–sample matrix of recurrently mutated genes, ranked by mutation frequency in the validation cohort of PCNSL patients (n = 82).

E: GISTIC2.0 results of significant recurrent focal amplifications (red) and deletions (blue). Genes affected by each focal event are annotated in the discovery cohort (n = 58). X-axis: plot of chromosomes; Y-axis: G score.

F: GISTIC2.0 results of significant recurrent focal amplifications (red) and deletions (blue). Genes affected by each focal event are annotated in the validation cohort (n = 82). X-axis: plot of chromosomes; Y-axis: G score.

G: The most significant contributions to the PCNSL genome in the discovery cohort (n = 58).

H: The most significant contributions to the PCNSL genome in the validation cohort (n = 82).

Related to Figure 2.

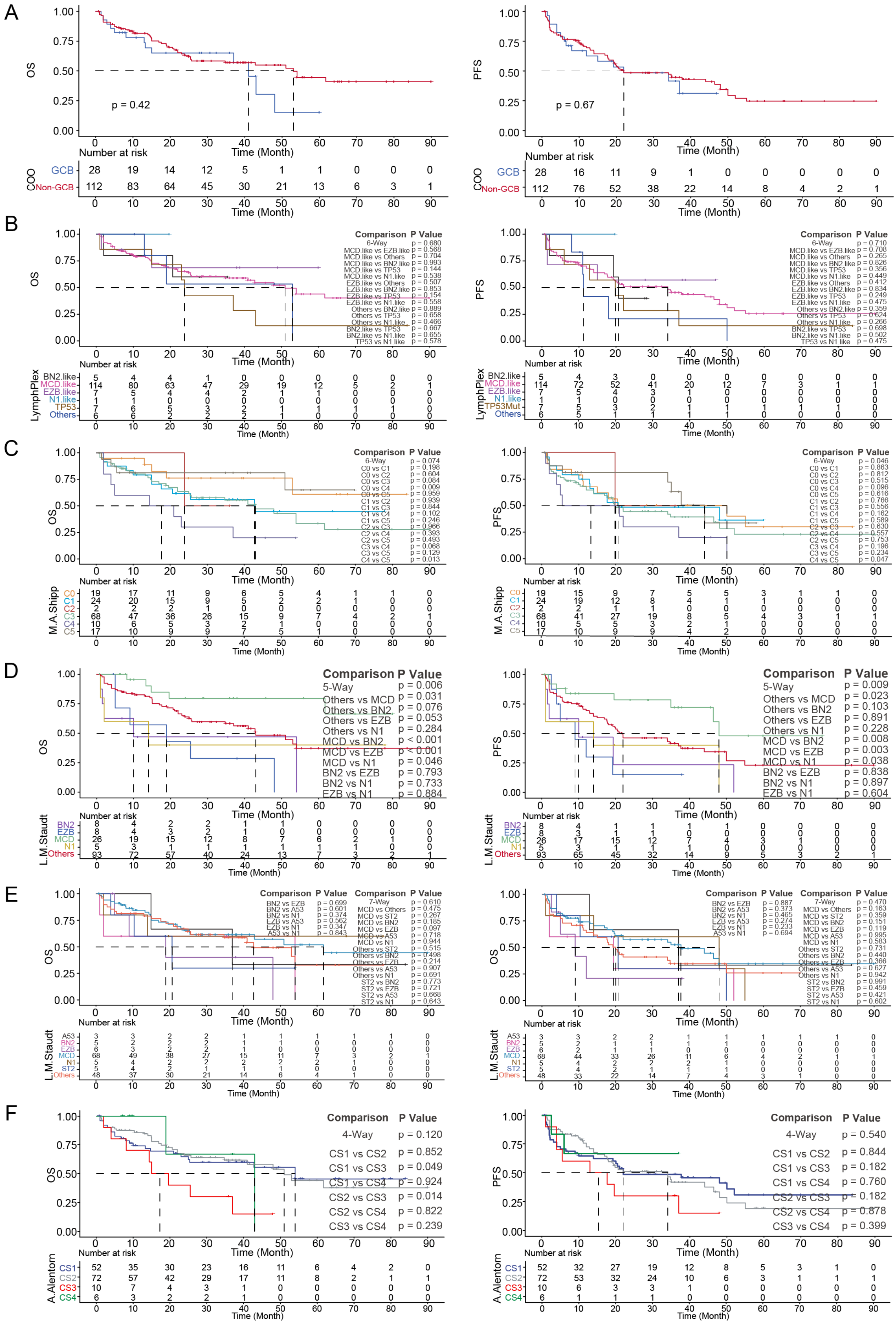

A
OS
p = 0.42
Number at risk
COO
GCB 28 19 14 12 5 1 1 0 0 0
Non-GCB 112 83 64 45 30 21 13 6 3 1
Time (Month)
PFS
p = 0.67
GCB 28 16 11 9 1 0 0 0 0 0
Non-GCB 112 76 52 38 22 14 8 4 2 1
B
LymphPlex
BN2.like
MCD.like
EZB.like
N1.like
TP53
Others
Comparison P Value
C
M.A.Shipp
C0
C1
C2
C3
C4
C5
D
Comparison P Value
5-Way p = 0.006
Others vs MCD p = 0.031
Others vs BN2 p = 0.076
Others vs EZB p = 0.053
Others vs N1 p = 0.284
MCD vs BN2 p < 0.001
MCD vs EZB p < 0.001
MCD vs N1 p = 0.046
BN2 vs EZB p = 0.793
BN2 vs N1 p = 0.733
EZB vs N1 p = 0.884
5-Way p = 0.009
Others vs MCD p = 0.023
Others vs BN2 p = 0.103
Others vs EZB p = 0.891
Others vs N1 p = 0.228
MCD vs BN2 p = 0.008
MCD vs EZB p = 0.003
MCD vs N1 p = 0.038
BN2 vs EZB p = 0.838
BN2 vs N1 p = 0.897
EZB vs N1 p = 0.604
L.M.Staudt
BN2
EZB
MCD
N1
Others
E
L.M.Staudt
A53
BN2
EZB
MCD
N1
ST2
Others
F
Comparison P Value
4-Way p = 0.120
CS1 vs CS2 p = 0.852
CS1 vs CS3 p = 0.049
CS1 vs CS4 p = 0.924
CS2 vs CS3 p = 0.014
CS2 vs CS4 p = 0.822
CS3 vs CS4 p = 0.239
4-Way p = 0.540
CS1 vs CS2 p = 0.844
CS1 vs CS3 p = 0.182
CS1 vs CS4 p = 0.760
CS2 vs CS3 p = 0.182
CS2 vs CS4 p = 0.878
CS3 vs CS4 p = 0.399
A.Alentorn
CS1 52 35 30 23 16 11 6 4 2 0
CS2 72 57 42 29 17 11 8 2 1 1
CS3 10 7 4 3 1 0 0 0 0 0
CS4 6 3 2 2 1 0 0 0 0 0
CS1 52 32 27 19 12 8 5 3 1 0
CS2 72 53 32 24 10 6 3 1 1 1
CS3 10 6 3 3 1 0 0 0 0 0
CS4 6 1 1 1 0 0 0 0 0 0

**Figure S6. The prognostic value of published molecular subtypes for progression-free survival (PFS) and overall survival (OS) in our Chinese PCNSL cohort**

A: Prognostic value of the DLBCL COO molecular subtype (GCB, non-GCB) for OS and PFS in our Chinese PCNSL cohort

B: The prognostic value of DLBCL LymphPlex molecular subtypes (BN2, MCD, EZB, N1, TP53, and others) for OS and PFS in our Chinese PCNSL cohort

C: The prognostic value of M.A. Shipp's team's DLBCL molecular subtypes (C0, C1, C2, C3, C4, and C5) for OS and PFS in our Chinese PCNSL cohort

D: The prognostic value of L.M. Staudt's team's DLBCL molecular subtypes (BN2, EZB, MCD, N1, and others) for OS and PFS in our Chinese PCNSL cohort

E: The prognostic value of L.M. Staudt's DLBCL molecular subtypes (A53, BN2, EZB, MCD, N1, ST2, and others) for OS and PFS in our Chinese PCNSL cohort.

F: Prognostic value of A. Alentorn team's PCNSL molecular subtypes (CS1, CS2, CS3, and CS4) for OS and PFS in our Chinese PCNSL cohort

Related to Figure 3.

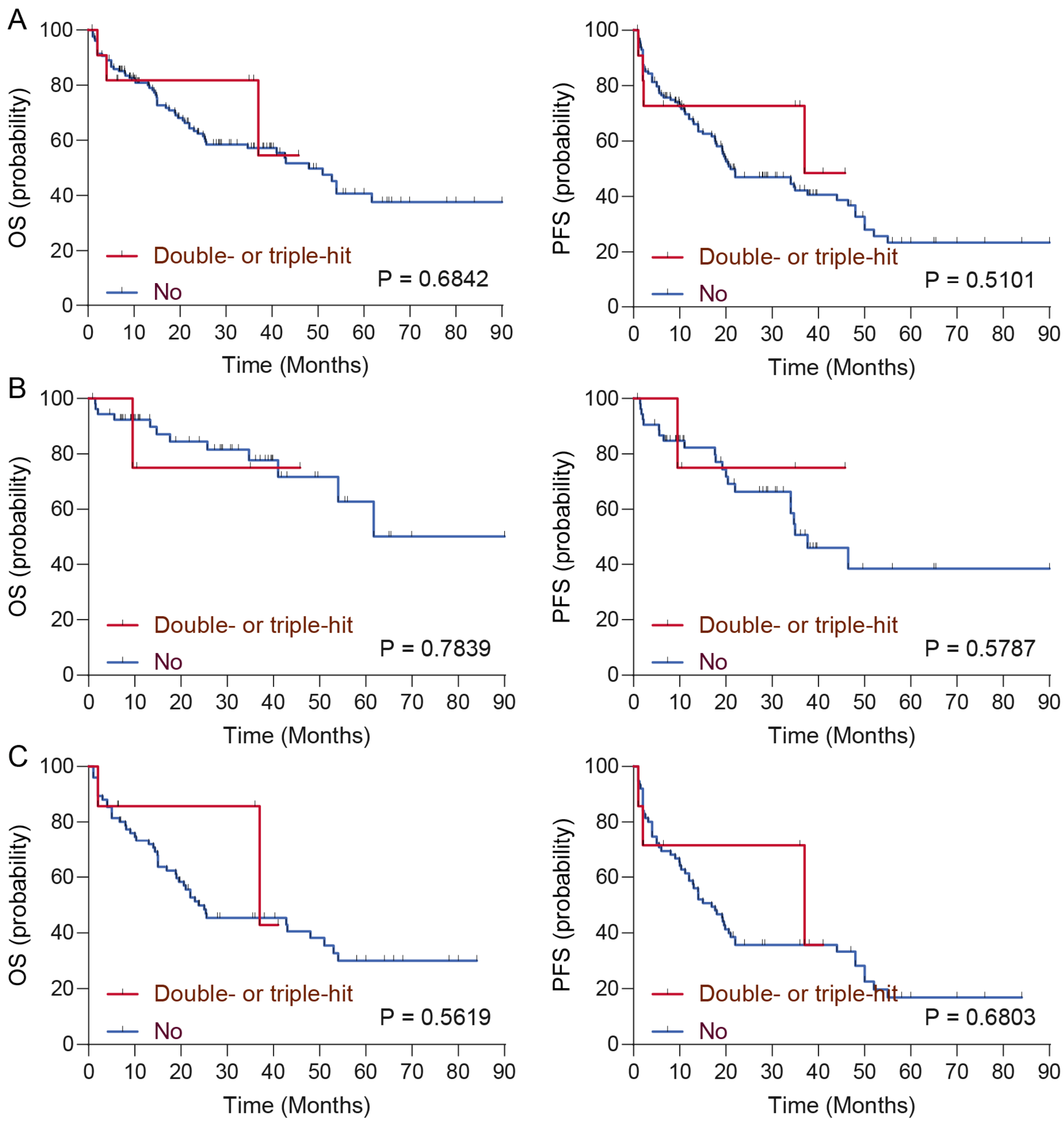


**Figure S7. The prognostic value of the double-hit gene expression signature subtypes for progression-free survival (PFS) and overall survival (OS) in our Chinese PCNSL cohort**

A: The prognostic value of double-hit gene expression signature subtypes (double or triple hit vs. others) for OS and PFS in our Chinese PCNSL discovery cohort (n = 58)

B: The prognostic value of double-hit gene expression signature subtypes (double or triple hit vs. others) for OS and PFS in our Chinese PCNSL validation cohort (n = 82)

C: The prognostic value of double-hit gene expression signature subtypes (double or triple hit vs. others) for OS and PFS in our Chinese PCNSL cohort (n = 140).

Related to Figure 3.

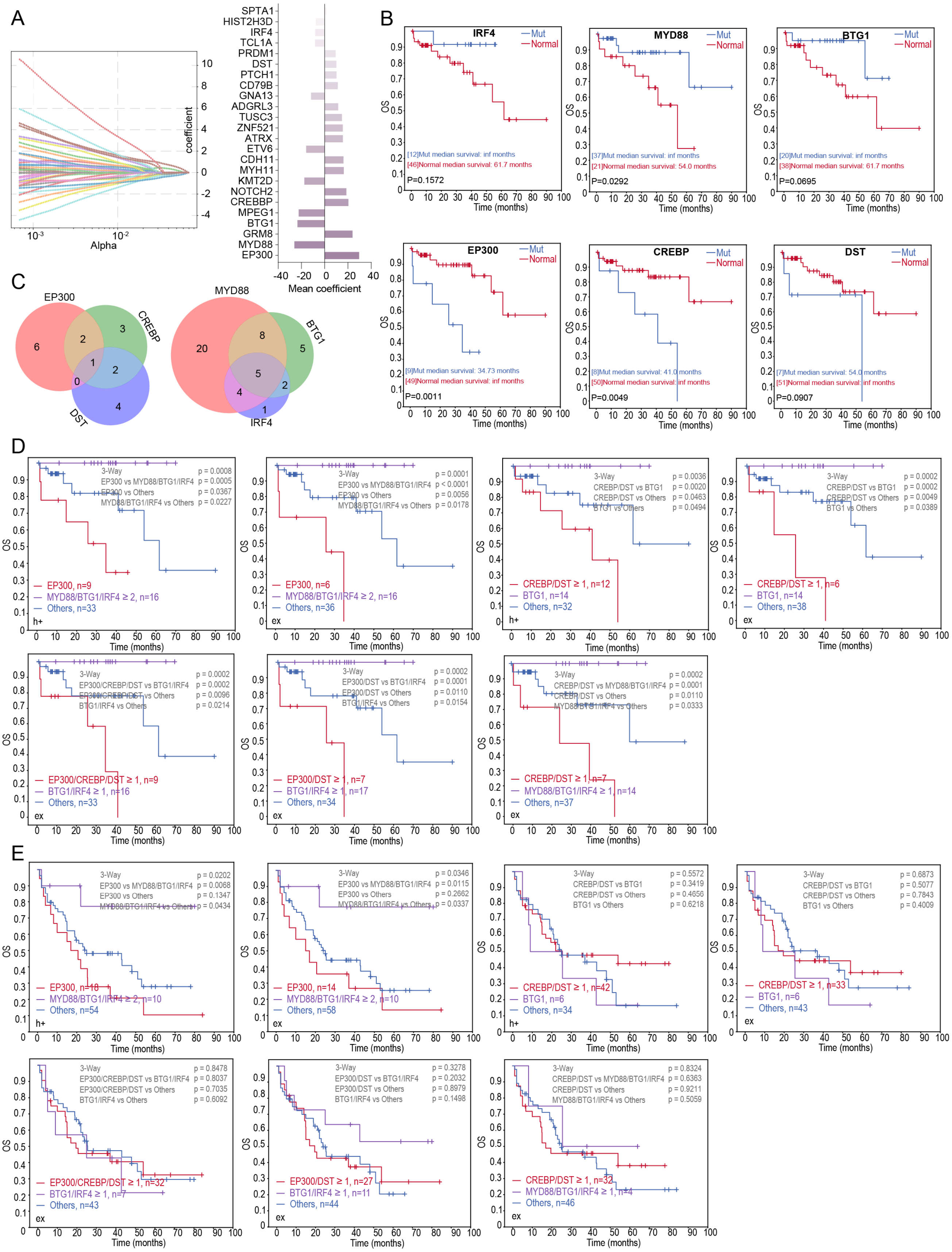

**Figure S8. Schematic of the typing strategies used to identify PCNSL molecular subtypes**

A: CoxNet survival analyses and least absolute shrinkage and selection operator (LASSO) regressions were employed to determine the importance score of each feature in the model, all of which were then ranked. This process was repeated 500 times, and all potential features were recorded. This is a demonstration of the aforementioned process.

B: Kaplan–Meier (K–M) analyses of the 6 identified features were conducted.

C: Venn diagram illustrating how these 6 features were randomly combined to form three types.

D: Seven candidate molecular subtype combinations were identified in the discovery cohort.

E: Seven candidate molecular subtype combinations were validated in the validation cohort.

Related to Figure 4.

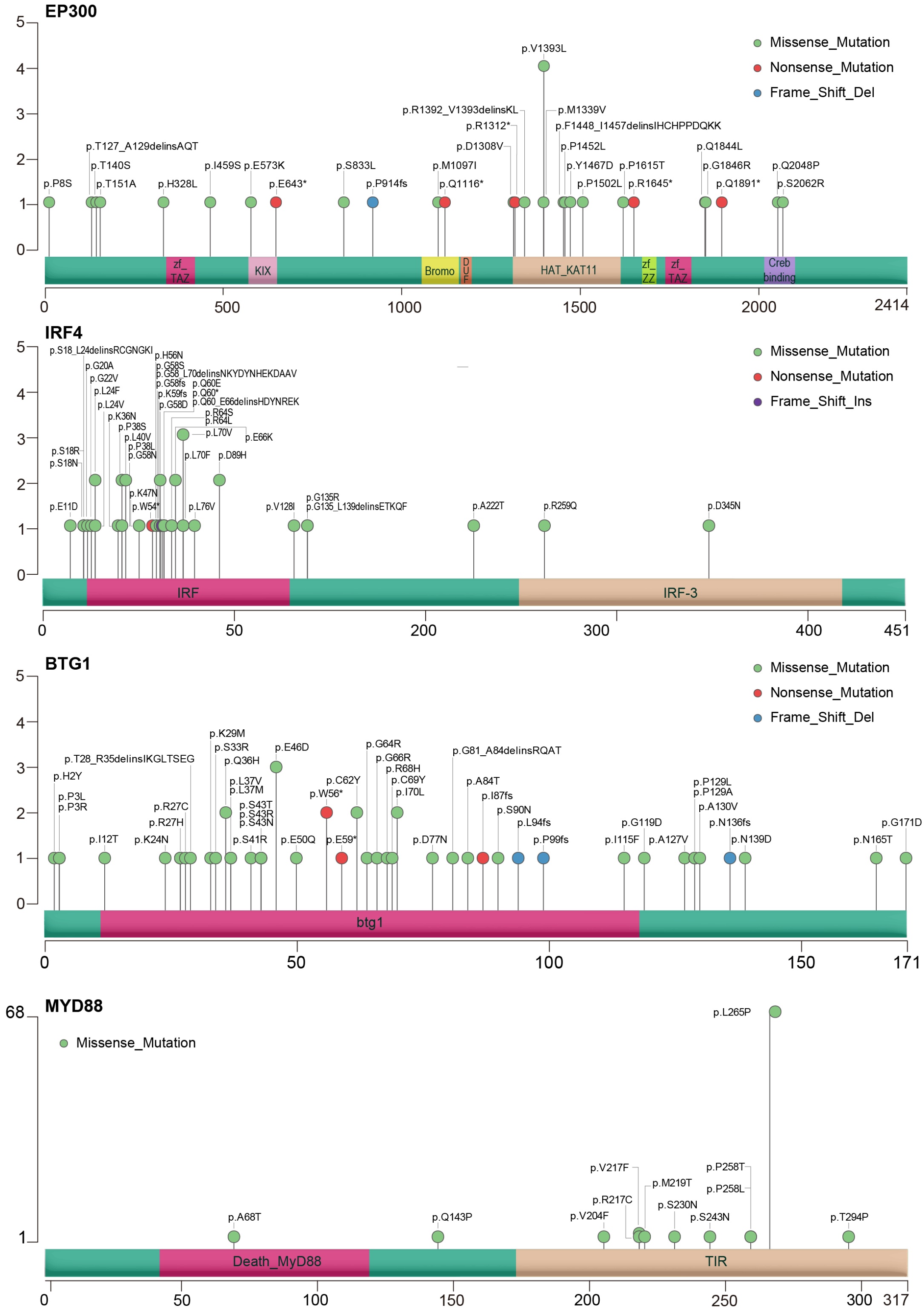


**Figure S9. Mutation diagrams (Lollipop Figures) for the EP300, IRF4, BTG1, and MYD88 genes**

For each gene, all nonsynonymous mutations within the functional domains of the respective protein were visualized via maftools.

Related to Figure 4.

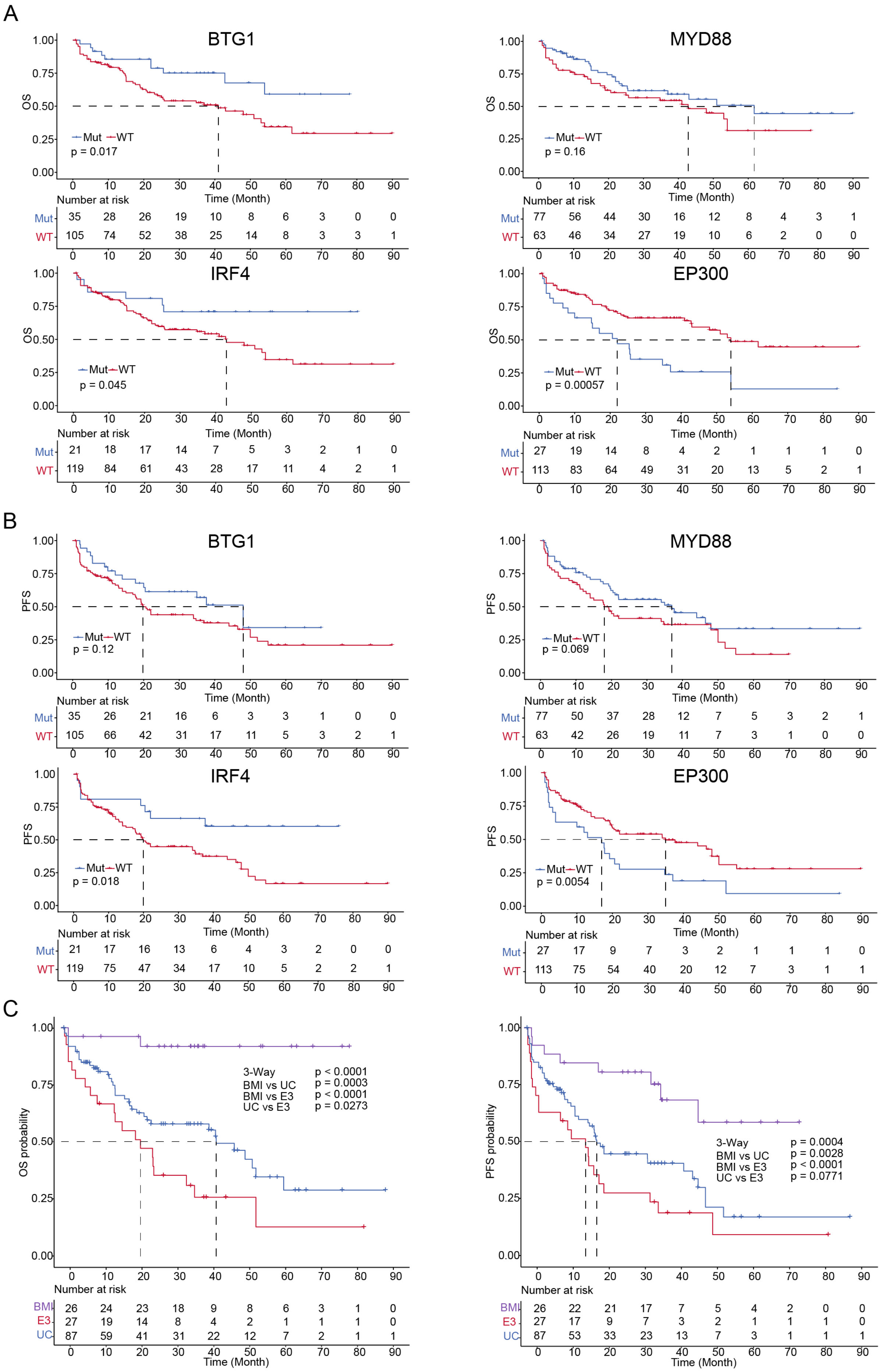
A
BTG1
OS
1.00
0.75
0.50
0.25
0.00
Mut WT
p = 0.017
Time (Month)
Number at risk
Mut 35 28 26 19 10 8 6 3 0 0
WT 105 74 52 38 25 14 8 3 3 1
0 10 20 30 40 50 60 70 80 90
MYD88
OS
Mut WT
p = 0.16
Time (Month)
Number at risk
Mut 77 56 44 30 16 12 8 4 3 1
WT 63 46 34 27 19 10 6 2 0 0
0 10 20 30 40 50 60 70 80 90
IRF4
OS
Mut WT
p = 0.045
Time (Month)
Number at risk
Mut 21 18 17 14 7 5 3 2 1 0
WT 119 84 61 43 28 17 11 4 2 1
0 10 20 30 40 50 60 70 80 90
EP300
OS
Mut WT
p = 0.00057
Time (Month)
Number at risk
Mut 27 19 14 8 4 2 1 1 1 0
WT 113 83 64 49 31 20 13 5 2 1
0 10 20 30 40 50 60 70 80 90
B
BTG1
PFS
Mut WT
p = 0.12
Time (Month)
Number at risk
Mut 35 26 21 16 6 3 3 1 0 0
WT 105 66 42 31 17 11 5 3 2 1
0 10 20 30 40 50 60 70 80 90
MYD88
PFS
Mut WT
p = 0.069
Time (Month)
Number at risk
Mut 77 50 37 28 12 7 5 3 2 1
WT 63 42 26 19 11 7 3 1 0 0
0 10 20 30 40 50 60 70 80 90
IRF4
PFS
Mut WT
p = 0.018
Time (Month)
Number at risk
Mut 21 17 16 13 6 4 3 2 0 0
WT 119 75 47 34 17 10 5 2 2 1
0 10 20 30 40 50 60 70 80 90
EP300
PFS
Mut WT
p = 0.0054
Time (Month)
Number at risk
Mut 27 17 9 7 3 2 1 1 1 0
WT 113 75 54 40 20 12 7 3 1 1
0 10 20 30 40 50 60 70 80 90
C
OS probability
3-Way p < 0.0001
BMI vs UC p = 0.0003
BMI vs E3 p < 0.0001
UC vs E3 p = 0.0273
Number at risk
BMI 26 24 23 18 9 8 6 3 1 0
E3 27 19 14 8 4 2 1 1 1 0
UC 87 59 41 31 22 12 7 2 1 1
0 10 20 30 40 50 60 70 80 90
Time (Month)
PFS probability
3-Way p = 0.0004
BMI vs UC p = 0.0028
BMI vs E3 p < 0.0001
UC vs E3 p = 0.0771
Number at risk
BMI 26 22 21 17 7 5 4 2 0 0
E3 27 17 9 7 3 2 1 1 1 0
UC 87 53 33 23 13 7 3 1 1 1
0 10 20 30 40 50 60 70 80 90
Time (Month)

**Figure S10. Kaplan–Meier analyses of the EP300, IRF4, BTG1, and MYD88 genes and molecular subtypes**

A: Prognostic value of mutations in the EP300, IRF4, BTG1, and MYD88 genes for OS.

B: Prognostic value of mutations in the EP300, IRF4, BTG1, and MYD88 genes for progression-free survival (PFS).

C: Kaplan–Meier estimates of overall survival (OS) and progression-free survival (PFS) among 140 PCNSL patients with each molecular subtype.

Related to Figure 4.

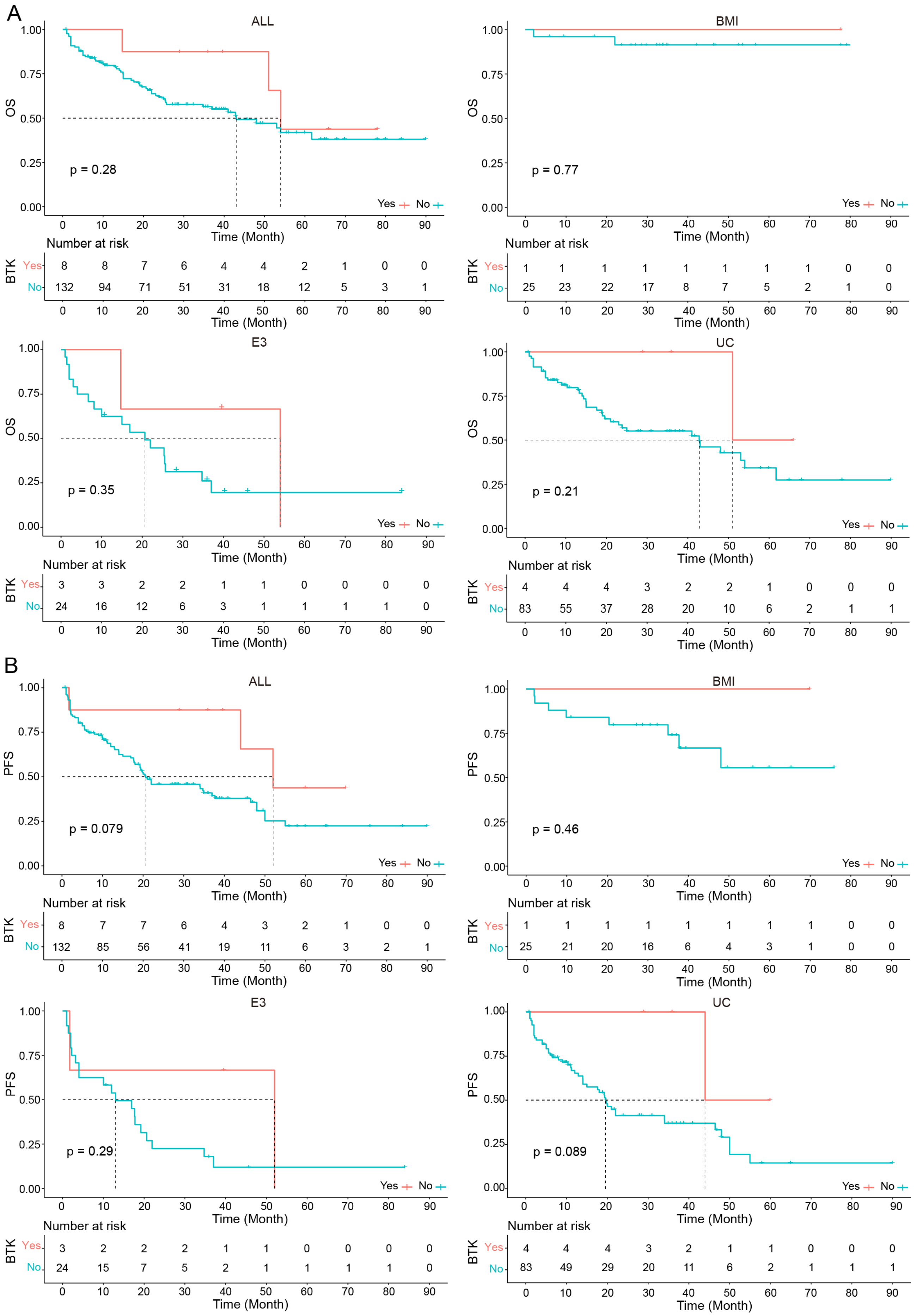

A
ALL
OS
p = 0.28
Yes No
Time (Month)
Number at risk
BTK Yes 8 8 7 6 4 4 2 1 0 0
No 132 94 71 51 31 18 12 5 3 1
BMI
p = 0.77
BTK Yes 1 1 1 1 1 1 1 1 0 0
No 25 23 22 17 8 7 5 2 1 0
E3
p = 0.35
BTK Yes 3 3 2 2 1 1 0 0 0 0
No 24 16 12 6 3 1 1 1 1 0
UC
p = 0.21
BTK Yes 4 4 4 3 2 2 1 0 0 0
No 83 55 37 28 20 10 6 2 1 1
B
ALL
PFS
p = 0.079
BTK Yes 8 7 7 6 4 3 2 1 0 0
No 132 85 56 41 19 11 6 3 2 1
BMI
p = 0.46
BTK Yes 1 1 1 1 1 1 1 1 0 0
No 25 21 20 16 6 4 3 1 0 0
E3
p = 0.29
BTK Yes 3 2 2 2 1 1 0 0 0 0
No 24 15 7 5 2 1 1 1 1 0
UC
p = 0.089
BTK Yes 4 4 4 3 2 1 1 0 0 0
No 83 49 29 20 11 6 2 1 1 1

**Figure S11. Kaplan–Meier estimates of the impact of BTK inhibitor use on prognosis**

A: The prognostic value of BTK inhibitor use for overall survival (OS) in all 140 PCNSL patients and patients with each molecular subtype.

B: Prognostic value of BTK inhibitor use for progression-free survival (PFS) in all 140 PCNSL patients and patients with each molecular subtype.

Related to Figure 4.

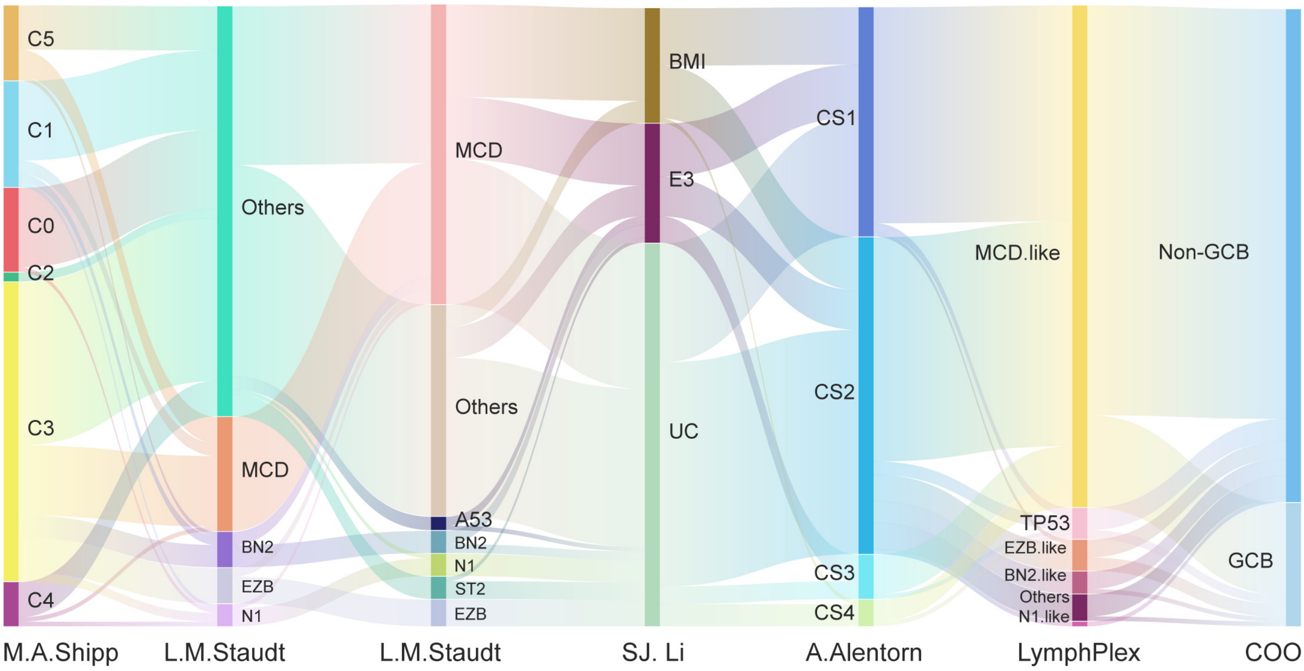


**Figure S12.** Sankey diagram illustrating the relationships between our PCNSL molecular subtypes and the published DLBCL and PCNSL molecular subtypes.

Related to Figure 4.

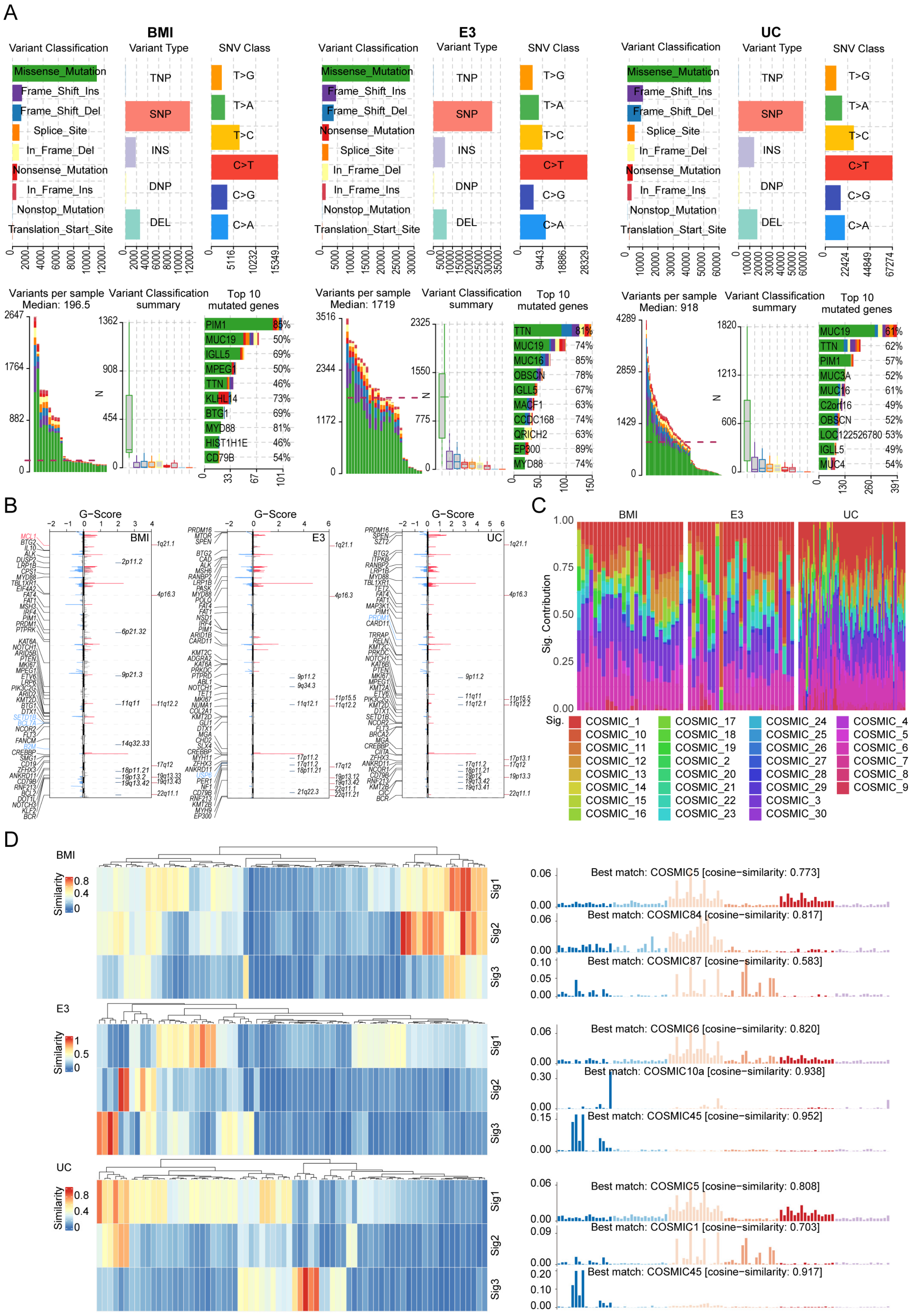
A
BMI
E3
UC
Variant Classification
Variant Type
SNV Class
Missense_Mutation
Frame_Shift_Ins
Frame_Shift_Del
Splice_Site
In_Frame_Del
Nonsense_Mutation
In_Frame_Ins
Nonstop_Mutation
Translation_Start_Site
TNP
SNP
INS
DNP
DEL
T>G
T>A
T>C
C>T
C>G
C>A
Variants per sample Median: 196.5
Variants per sample Median: 1719
Variants per sample Median: 918
Variant Classification summary
Top 10 mutated genes
B
G-Score
C
Sig. Contribution
D
Similarity
Sig1
Sig2
Sig3
Best match: COSMIC5 [cosine−similarity: 0.773]
Best match: COSMIC84 [cosine−similarity: 0.817]
Best match: COSMIC87 [cosine−similarity: 0.583]
Best match: COSMIC6 [cosine−similarity: 0.820]
Best match: COSMIC10a [cosine−similarity: 0.938]
Best match: COSMIC45 [cosine−similarity: 0.952]
Best match: COSMIC5 [cosine−similarity: 0.808]
Best match: COSMIC1 [cosine−similarity: 0.703]
Best match: COSMIC45 [cosine−similarity: 0.917]

**Figure S13. Mutation profiles and signatures across the three MSs**

A. Variant classification, type, and single-nucleotide variation (SNV) class across the E3, BMI, and UC subtypes.

B. GISTIC2.0 results of significant recurrent focal amplifications (red) and deletions (blue). Genes affected by each focal event are annotated. X-axis: plot of chromosomes; Y-axis: G score.

C. Contributions of mutational signatures across the E3, BMI, and UC subtypes.

D. The most significant contributions to the PCNSL genome across the E3, BMI, and UC subtypes.

Related to Figure 5.

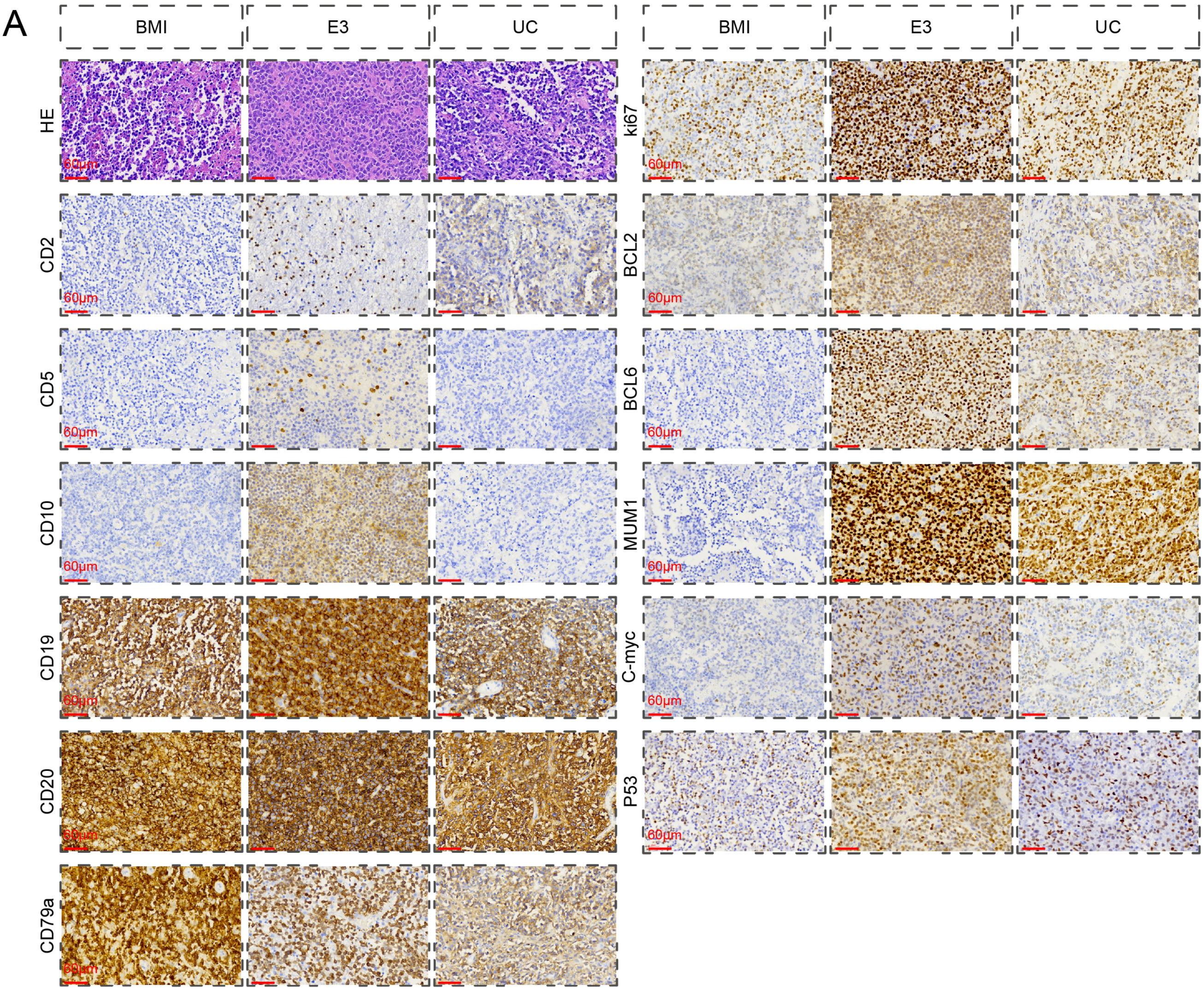


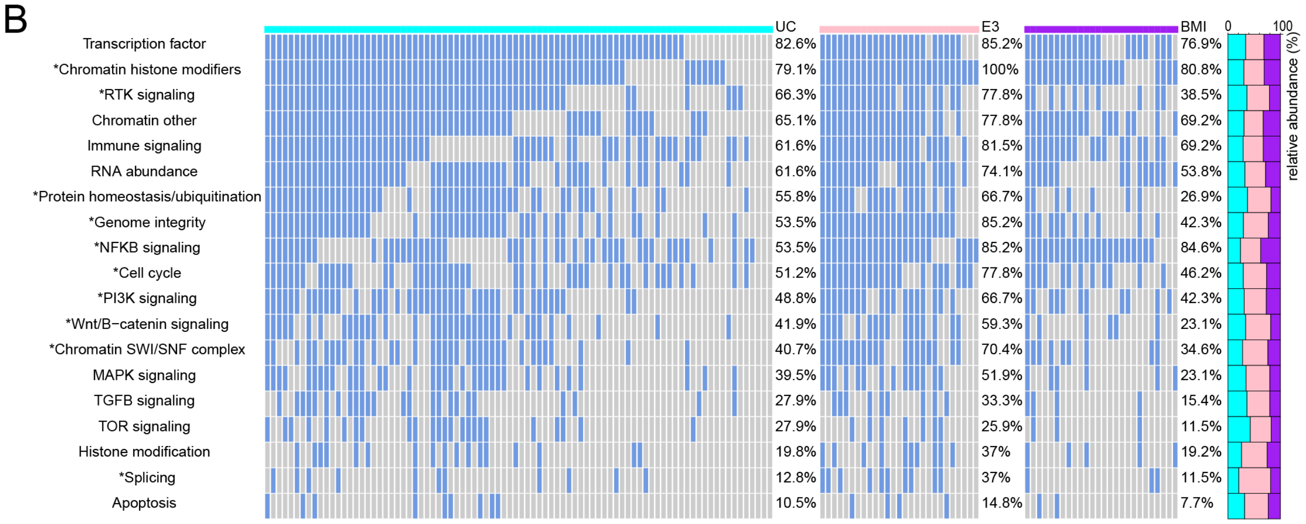


**Figure S14.** Hematoxylin–eosin staining and immunohistochemistry (A) for CD2, CD5, CD10, CD19, CD20, CD79a, Ki-67, BCL2, BCL6, MUM1, C-Myc, and p53, and oncogene pathways (B) in samples from different PCNSL subtypes

Related to Figure 5.

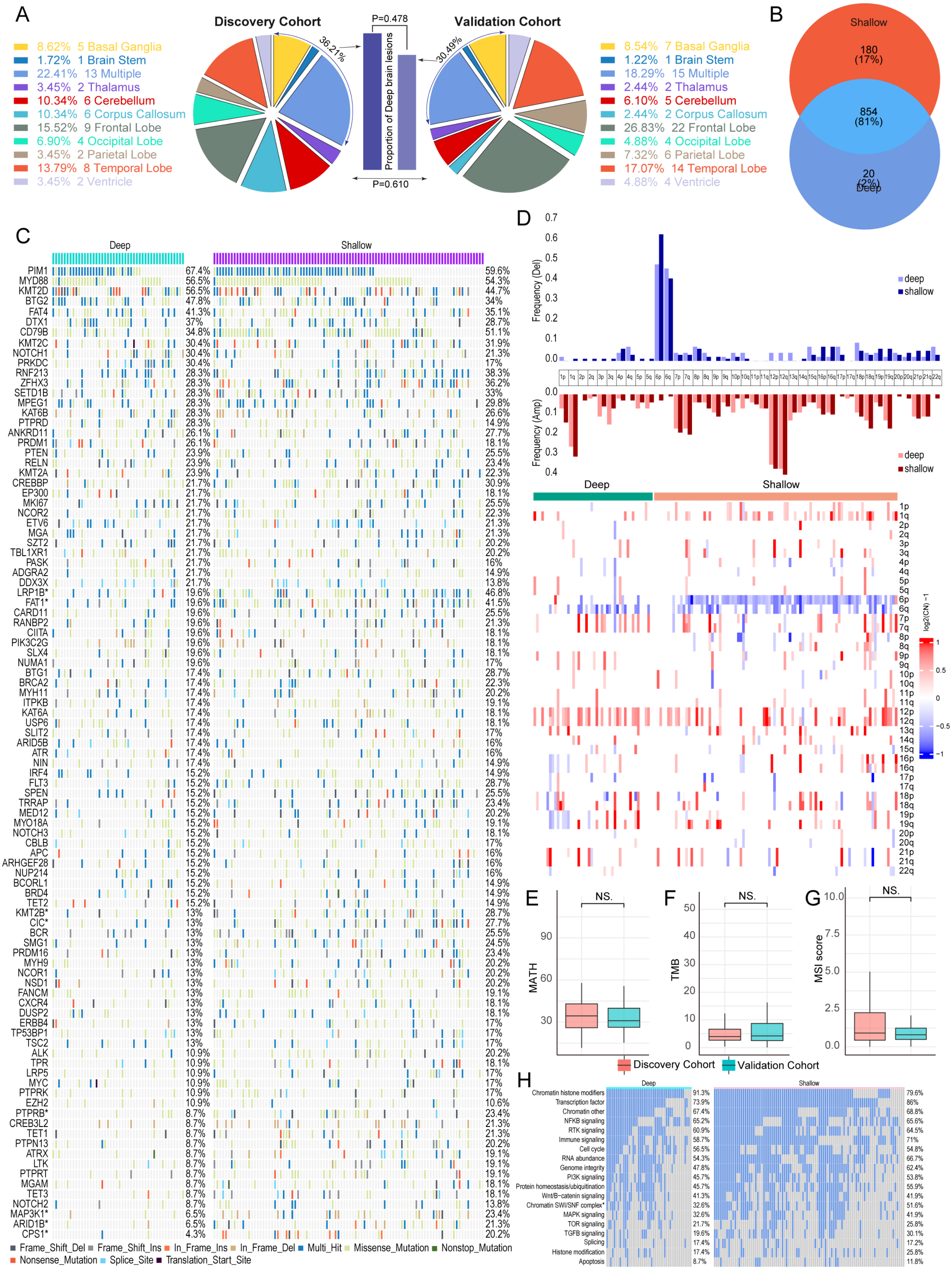


**Figure S15. Mutational profiles of deep brain and shallow brain tumors in PCNSL**

A: Difference in tumor sites in the discovery versus validation cohorts.

B: Venn diagram of unique and shared mutations in deep brain and shallow brain tumors.

C: The number and frequency of recurrent mutations, along with a gene-sample matrix of recurrently mutated genes in PCNSL patients between deep brain and shallow brain tumors, are presented. The relative abundance across the molecular subtypes is displayed on the right.

D: Arm-level copy number alterations between deep brain and shallow brain tumors are displayed. The frequencies of amplifications and deletions were compared.

E: The levels of mutant-allele tumor heterogeneity (MATH) were compared between deep brain and shallow brain tumors.

F: The tumor mutational burden (TMB) was compared between deep brain and shallow brain tumors.

G: The levels of microsatellite instability (MSI) were compared between deep brain and shallow brain tumors.

H: Pathways affected by oncogenes in PCNSL patients between deep brain and shallow brain tumors.

The chi-squared test and one-way ANOVA were used. $^{*}P < 0.05$, $^{ns}P > 0.05$

Related to Figure 6.
